# Clustering of adverse health and educational outcomes in adolescence following early childhood poverty: implications for UK’s ‘levelling up’ agenda

**DOI:** 10.1101/2022.08.11.22278671

**Authors:** Aase Villadsen, Miqdad Asaria, Ieva Skarda, George B. Ploubidis, Mark Mon Williams, Eric John Brunner, Richard Cookson

## Abstract

**Background:** Early childhood poverty is associated with poorer health and educational outcomes in adolescence. However, there is limited evidence about the clustering of these adverse outcomes by income group.

**Methods:** We analysed five outcomes at age 17 known to limit life chances – psychological distress, self-assessed ill health, smoking, obesity, and poor educational achievement – using data from the longitudinal UK Millennium Cohort Study (N=15,245). We compared how single and multiple outcomes were distributed across quintiles of household income in early childhood (0-5 years) and modelled the maximum potential benefit of tackling the income gradient in these outcomes.

**Findings:** Children from the poorest households were 12.7(95% CI 6.4-25.1) times more likely than those from the richest to experience four or five adverse adolescent outcomes, with poor educational achievement and smoking showing the largest single risk ratios–4.6(95% CI: 4.2-5.0) and 3.6(95% CI 3.0-4.2), respectively. We modelled hypothetical absolute and relative poverty elimination scenarios, as well as an income inequality elimination scenario, and found these would yield maximum reductions in multiple adolescent adversity of 5%, 30%, and 80% respectively.

**Interpretation:** Early childhood poverty is more strongly correlated with multiple adolescent adversity than any single adverse outcome. Reducing absolute poverty alone is not sufficient to eliminate the life-long burden of multiple adversity, which disproportionately impacts children across the bottom three-fifths of the income distribution. An ambitious levelling up agenda needs co-ordinated multi-agency action to tackle the complex interacting factors generating the steep social gradient in multiple adolescent adversity.

## INTRODUCTION

The child poverty rate is returning to its mid-1990 level (33%), with almost a third of children living below the poverty line in 2019–20 after housing costs (31%, 4.3□million).^1^ Children in lone parent families and from black and minority ethnic groups are particularly likely to live in poverty.^2^ The COVID-19 pandemic and cost of living crisis are amplifying these trends. In the absence of effective policy intervention, rates of child poverty are set to rise further, with inevitable damage to the education, physical and mental health, as well as life chances, of those exposed to early material deprivation.

Socioeconomic gradients in health and development are initiated in early childhood.^3-5^ Long-term associations between lower household income in childhood and worse performance on a range of educational outcomes, health outcomes and health behaviours have been observed in recent cohorts in the UK and other high income countries.^6 7^ These adolescent outcomes in turn drive adult outcomes such as employment status, income and wealth, life satisfaction, and lifetime experiences of physical and mental illness.^8 9^

The problem of inequalities is recognised at government level, and the UK government response is currently encapsulated in the political slogan ‘Levelling Up’, which refers broadly to reducing inequalities in life chances and health.^10^

Existing literature on the effects of early childhood poverty examines single adverse outcomes in adolescence separately.^11-22^ Further, studies of inequality in the cohort born after the Millennium (Generation Z) are sparse. In the present economic and welfare policy context, it is important to understand in detail the experience of those coming of age at present and to consider the implications for population health and policy in coming decades.

We use data from all seven waves of the Millennium Cohort Study (MCS), the most recent nationally representative longitudinal study of young people available in the UK. We not only study key adolescent health and educational outcomes separately but also examine how these outcomes cluster according to childhood household income. Based on observed distributions, we also calculate counterfactual outcome prevalence according to three ‘levelling-up’ scenarios. Our objectives are, first, to describe the association between household income in early childhood (ages 0-5 years) and physical health, psychological distress, smoking behaviour, obesity, and educational outcomes at age 17 years. We look at the patterning and clustering of these five outcomes by income quintile. Second, we examine the potential benefits of altering the income distribution among households with young children in terms of reducing the population burden of adverse adolescent outcomes.

## METHODS

### Data

The Millennium Cohort Study (MCS) is a longitudinal study that has followed around 19,000 individuals born in the UK around the millennium (Sep 2000-Jan 2002). A detailed description of the MCS - including sampling frame and strategy, and inclusion criteria - is provide elsewhere.^23^ The analytical sample used in the current study includes 15,245 participants. See the ‘multiple imputations and weighting’ section in the supplementary material for how this sample was derived. Ethical approval was obtained for each of the survey sweeps, with informed consent obtained from the parents of cohort members up to age 14 and from age 17 onwards also from the cohort members themselves.

### Measures

*Household income in early childhood* was reported by the main parent at age 9 months, age 3 and 5 years (data collected in 2001-2006). Banded responses were used to impute continuous income, which was then equivalised to take account of household size and composition using modified OECD scales.^24^ Household income was averaged across these first three waves of MCS and transformed into income quintile groups for use in this study.

Adverse health and social outcomes in adolescence were captured at age 17 (data collected in 2018) based on five measures that we constructed by dichotomising responses to questions from the seventh wave of the MCS. The first measure was *psychological distress* which was self-reported by the cohort members using the six-item Kessler scale (K6). We used the clinically determined threshold (13 and above) as indicating the presence of psychological distress.^25^ The second measure was *self-rated health*, which was assessed through the question “How would you describe your health generally?”. We classified responses of ‘excellent’, ‘very good’, and ‘good’ as being in good health, and responses of ‘fair’ and ‘poor’ classified as being in ‘poor’ health. The third measure was *obesity*, based on the adolescents’ weight and height taken in the home by the interviewer, and classified using the obesity threshold from the British 1990 (UK90) growth reference chart for children.^26^ The fourth measure was *regular cigarette smoking* (excluding e-cigarettes) which was self-reported smoking of more than six cigarettes a week. The fifth measure was *poor academic achievement* based of self-reported exam results at the end of secondary school. In England, Wales and Northern Ireland these were GCSE results, and in Scotland results of N4 and N5. Poor academic attainment was classified as not achieving five or more GCSEs (including maths and English) graded C or above, or five or more N5s including maths and English graded D or above.

To examine the clustering of multiple outcomes we computed an *index of multiple adverse adolescent outcomes* by counting how many negative outcomes were present for each participant (range 0-5). We created a categorical variable that collapsed those with 4 and 5 adverse outcomes “0” (no adverse outcomes) “1” (precisely 1 adverse outcome), “2”, “3” and “4 or more”) as very few participants had five adverse outcomes.

Sex and ethnicity of cohort members, which were used in this study as moderators, were ascertained from the main parent during the initial survey.

### Analyses

We cross-tabulated childhood household income quintile groups and our index of multiple adverse adolescent outcomes at age 17 in the form of contingency tables showing the count in each cell alongside the row and column percentages. Our first research aim was to compare the strength of association between early childhood income and adverse adolescent outcome. To do this, we compared the proportions of respondents at each level of our index of adverse adolescent outcomes in the highest income quintile group to the proportions in each of the other income quintile groups. We used the *csi* function in Stata to calculate risk ratios and their 95% confidence intervals. We also calculated estimates for each single outcome separately. For the index of multiple adversities we stratified results by sex and by ethnicity (with interactions testing for any difference), which we report in the supplemental material.

Our second research aim was to do simple calculation about how far adverse outcomes at age 17 could hypothetically be reduced by levelling-up incomes from lower-income to higher-income groups. We used a population attributable fraction approach to examine the role of low income as a risk factor and calculated the impacts of three hypothetical “levelling up” scenarios on reducing adverse adolescent outcomes. In the first scenario we levelled up the respondents in the lowest income quintile group to the same level of adverse adolescent outcomes as those in second lowest income quintile group – we label this the *absolute poverty elimination* scenario. The second scenario was more ambitious and shifted the bottom two income groups to the middle income group – we label this the *relative poverty elimination* scenario. This final scenario was even more ambitious, and involved levelling up the whole population to the same level of adverse adolescent outcomes as those in the highest income quintile – we labelled this the *inequality elimination* scenario.

We did not include covariates when examining the association between childhood income and adverse adolescent outcomes. First, we did not want to over-adjust for mediating variables such as adverse experiences that may lie on the causal pathway between early childhood disadvantage and adolescent outcomes. Second, we wanted to use income as a proxy for general social disadvantage in childhood, rather than to focus on income specifically and independently of intersecting markers of social disadvantage.

Multiple imputations and weight were used to deal with attrition over time, and weight were also used to adjust for the complex initial sampling design (see supplemental material for details).

All analyses were carried out using STATA version 17.^27^

### Role of the funding source

The funders of the study had no role in study design, data collection, data analysis, data interpretation, or writing of the report.

## RESULTS

The main characteristics of our MCS participants (N=15,245) are reported in Table S1 (supplemental material). Children from households in the lowest income quintile group had an average weekly household income in childhood of £117 as compared to those from the highest group who had an average weekly household income in childhood of £644. On average, 14.7% of the age 17 sample experienced psychological distress, 7.6% reported poor health, 10.4% were regular smokers, 23.3% were obese, and 37.0% achieved poor academic outcomes. In terms of the clustering of adverse adolescent outcomes, 40.8% had none, 34.9% had one, 16.9% had two, 5.8% had three, and 1.7% had four or more. The total number of individuals with four or more adverse outcomes at age 17 in our sample was only 255, but when translated to the whole UK population that represents 12,121 individuals per year.

When looking separately at each of the five adverse outcomes at age 17 (Figure 1), the highest prevalence of any single adverse outcome among any group was poor academic achievement in the poorest quintile group of 63.6% [95% CI 60.8-66.3]. Relative differences between lowest and highest household income quintiles were largest for poor academic achievement (RR=4.6, 95% CI 4.2-5.0) and smoking (RR=3.6, 95% CI 3.3-4.2), and lowest for psychological distress (RR=1.5, 95% CI 1.3-1.7). See supplemental material Table S5 for formal testing of differences between all income groups. Cross-tabulations of childhood household income quintile groups against separate and multiple adverse adolescent outcomes are in supplemental material Tables S2 and S3, respectively.

**Figure 1:**
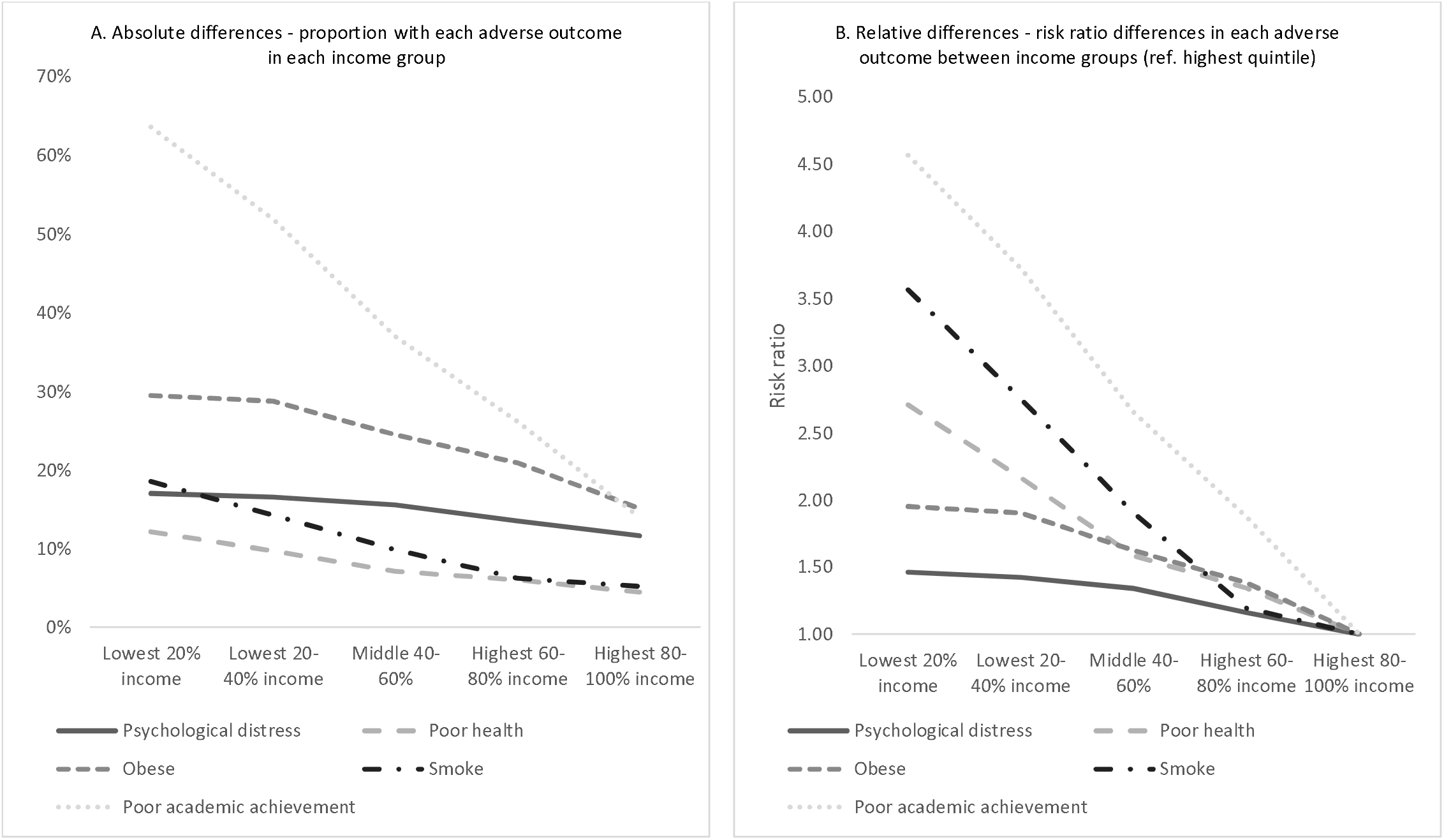
Five adverse outcomes at age 17 by early childhood income (N=15,245) Note: Panel A based row percentage in Table S2 (supplemental), Panel B based on Table S5 (supplemental). Refer to these tables for confidence intervals of estimates.

The clustering of adverse outcomes is shown in Figure 2 (see also cross-tabulations in Table S3, supplemental material). Inequalities based on childhood income were larger the more adverse outcomes were experienced in adolescence. Relative inequality between the lowest and highest income quintile groups was modest for one adverse outcome (RR=1.4, 95% CI 1.3-1.5), increasing in magnitude for two (RR=4.0, 95% CI 3.5-4.6), and for three (RR=4.5, 95% CI 3.5-5.7), and highest for four or five adverse outcomes (RR=12.7, 95% CI 6.4-25.1). The confidence intervals are wide for this estimate, because within the highest income quintile the number of individuals with four or five adverse group is extremely small – only 9 individuals in our sample, representing 0.3% of the highest income group, compared with 89 individuals within the lowest income group, representing 3.3% of that group (Table S3 in supplement). Risk ratio comparisons between the highest childhood household income quintile and all other income quintiles are reported in supplemental Table S6.

**Figure 2:**
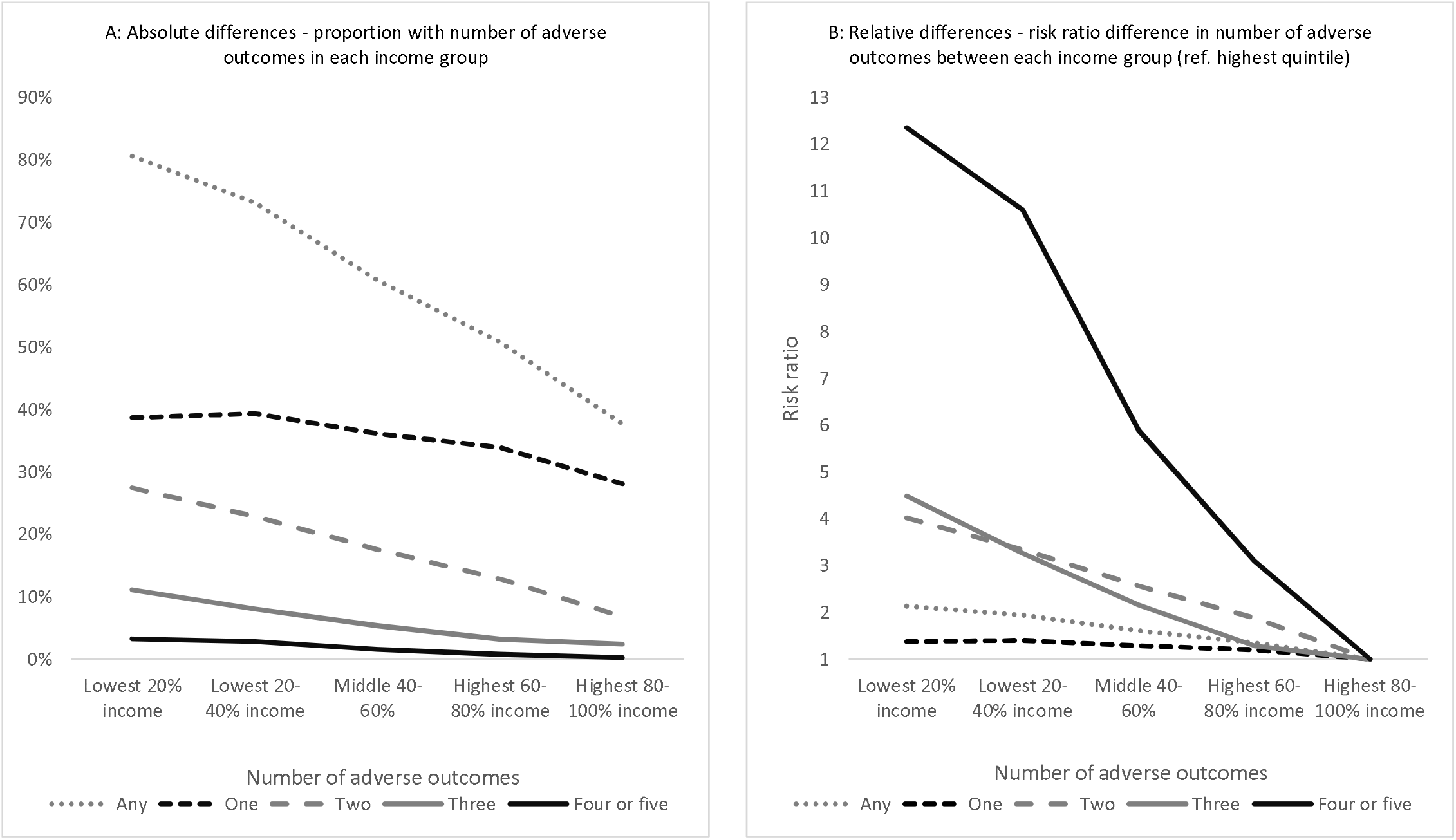
Number of adverse outcomes at age 17 by early childhood income (N=15,245) Note: Panel A based on row percentages Table S3 (supplemental), Panel B based on Table S6 (supplemental). Refer to these tables for confidence intervals of estimates.

In additional analyses we find little difference in the patterning of inequality in adverse adolescent outcomes by childhood household income by sex (supplemental material Figure S1, Figure S2, and Table S7). We find that inequalities are slightly greater (borderline significant interaction) for white cohort members than for cohort members from non-White ethnic groups in terms of having three or more adverse outcomes, suggesting that there may be some protective effects of non-white ethnicity (supplemental material Figure S3, Figure S4 and Table S8). Because Scottish school leaving results are measured differently to the other UK nations, we also carried analyses with the exclusion of Scotland, but found that the results are nearly identical to those using all four nations (supplemental material Figure S5 and S6).

The results of modelling the three levelling up scenarios are presented in Figure 3 (see also supplemental Table S2 and Table S3 on which results are based). Under the *absolute poverty elimination* scenario, we would reduce the total prevalence of adolescent psychological distress by 0.6%, self-reported poor health by 5.6%, obesity by 0.5%; regular smoking by 7.1% and poor academic achievement by 5.6%, under the *relative poverty elimination* scenario, we would reduce the total prevalence of psychological distress by 2.9%, self-reported poor health by 17.9%, obesity by 7.1%, regular smoking by 22.5% and poor academic achievement by 20.2%, whilst under the *inequality elimination* scenario we would reduce the total prevalence of psychological distress by 20.8%, self-reported poor health by 41.4%, obesity by 35.1%, regular smoking by 50.0% and poor academic achievement by 62.3%.

**Figure 3:**
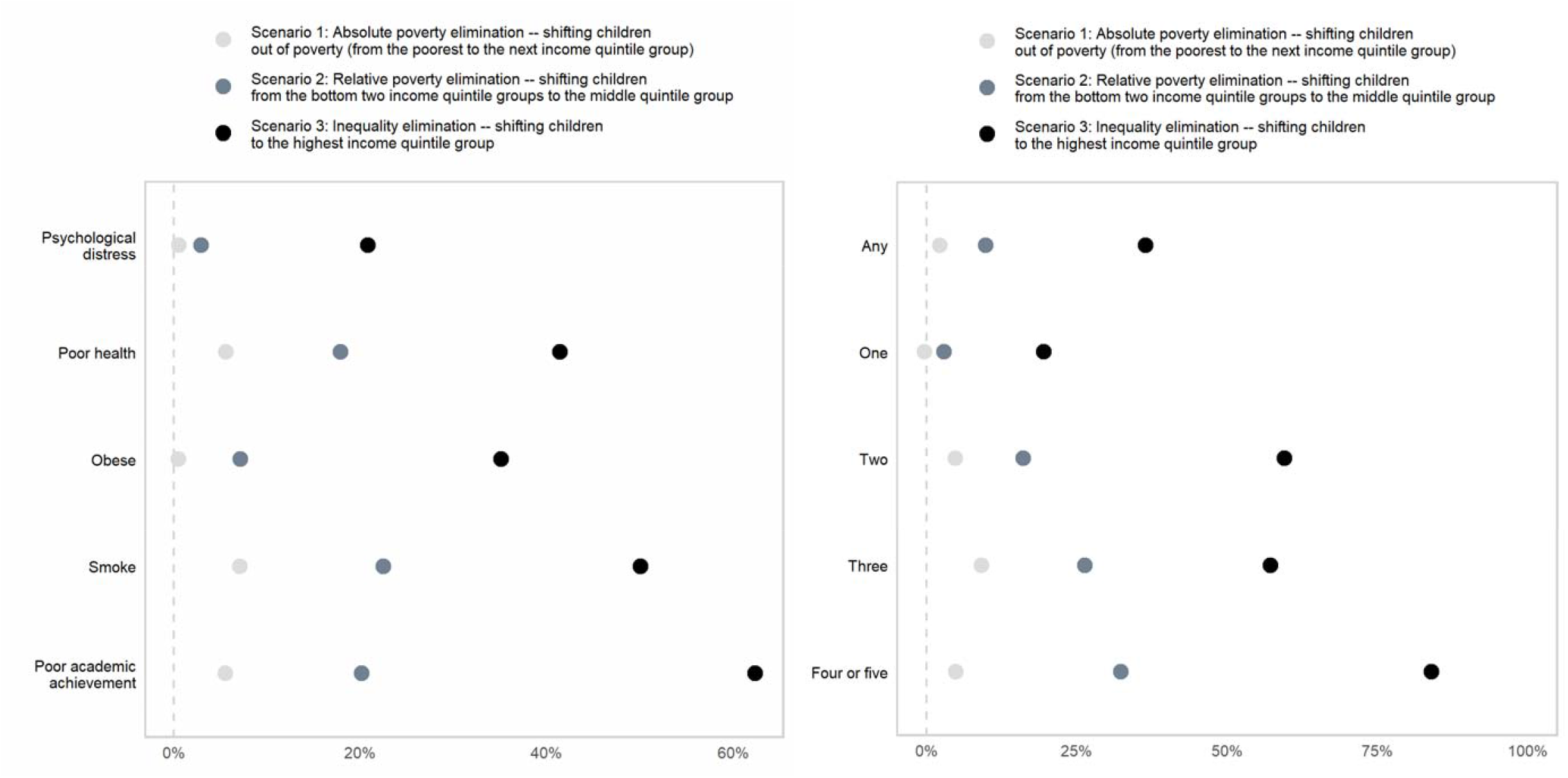
Proportional reduction in adverse outcomes from three hypothetical levelling up scenarios (N=15,245) Note: The proportional reductions are calculated relative to the baseline based on column percentage results in Table S2 and Table S3. Our scenario labels use convenient approximations to absolute and relative poverty based on the five income quintile groups used in our analysis. In the UK, the relative poverty line is officially defined as 60% of median income. Scenario 2 would therefore do more than enough to eliminate relative poverty based on the official definition. Confidence intervals of the estimates are shown in Table S9 (Supplemental)

In terms of the index of multiple adverse outcomes, we see that under the *absolute poverty elimination* scenario the proportion of adolescents having only one adverse outcome would increase by 0.3% (as some people who would have had more than one adverse outcome now have only one adverse outcome), the proportion having two adverse outcomes would reduce by 4.8%, the proportion having three adverse outcomes would reduce by 9.2%, and the proportion having four or five adverse outcomes would reduce by 4.9%. Under the *relative poverty elimination* scenario, the proportion of adolescents having only one adverse outcome would reduce by 3%, the proportion having two adverse outcomes would be reduced by 16.1%, the proportion having three adverse outcomes would reduce by 26.4%, and the proportion having four or five adverse outcomes would reduce by 32.3%. Under the *inequality elimination* scenario, the proportion of adolescents having one adverse outcome would reduce by 19.5%, the proportion having two adverse outcomes would reduce by 59.5%, the proportion having three adverse outcomes would reduce by 57.2%, and the proportion of people with four or five adverse outcomes would reduce by 83.9%.

Shown in Supplemental Table S9 are estimates of reduction in term of the absolute number of individuals in the current MCS sample as well as on a national level for each of the scenarios. Under the three scenarios, the simulated total annual reductions in the number of individuals in the UK with four or five adverse outcomes at age 17 are 597 (95% CI -2,879 to 4,074), 3,920 (-323 to 8,164) and 10,174 (95% CI: 5,718-14,631) per year, respectively.

## DISCUSSION

### Principal findings

Cohort members from the poorest early childhood household income quintile were almost 13 times more likely than those from the richest to experience multiple adolescent adversity involving four or more adverse adolescent outcomes. Only a small proportion experience four or more adverse adolescent outcomes – 1.67% in our sample – but when translated to the whole UK population that adds up to a total of about 12,121 additional adolescents per year leaving school with extremely poor life chances, representing a substantial and cumulative long-term burden of human and financial cost to society.

Hypothetical elimination of the early childhood income gradient could reduce this burden of multiple adversity by a maximum of eighty percent, and eliminating relative poverty in early childhood by shifting the bottom two income quintiles to the middle could reduce the burden by a maximum of thirty percent. However, levelling-up the income of the poorest children to the next poorest would only yield a maximum five percent reduction in multiple adolescent adversity involving four or more adverse outcomes.

### Comparison to previous research

Previous existing longitudinal studies have consistently shown that a range of adverse child and adolescent outcomes are patterned according to early childhood household income, with those growing up in the poorest families being much more likely to experience negative outcomes such as poor mental health, poor physical health, smoking, obesity, and poor academic achievement.^11-22^ The examination of each of our five single adverse outcomes confirm these previous findings.

We add to existing studies by showing that experiencing multiple adolescent adversity is much more strongly associated with socioeconomic disadvantage in early childhood than any single adverse outcome. We also contribute by providing estimates of the potential gains of alternative early childhood levelling-up strategies in terms of reducing the prevalence of health and educational adversity in adolescence. Previous studies have discussed the potential health and educational benefits of eliminating early childhood poverty, without providing quantitative estimates. We have attempted to provide upper bounds on various scenarios to tackle the income gradient in adverse adolescent outcomes.

### Strengths and limitations

The major strength of the study is its use of MCS, a representative population based longitudinal survey of those born in 2000. The richness of the dataset allowed us to explore a range of different outcomes in adolescence and the clustering of these outcomes in relation to childhood household income. In contrast, most previous studies on health inequalities have tended to focus on single outcomes and often rely on cross-sectional rather than longitudinal data.

Our study has a number of important limitations. In terms of the data, with the exception of obesity, we relied on self-reported measures for adverse outcomes. Whilst we have no reason to believe that any biases in self-reported values would be patterned by childhood household income this was not possible to verify and may have impacted our findings. In terms of methodology, our study identifies individual level associations between childhood household income and adverse adolescent outcomes. However, it does not identify the causal impact of changes in childhood household income, which is likely to be smaller than we estimate in our hypothetical reduction scenarios due to confounding factors (e.g. parental socio-emotional problems) that increase the risk of both early childhood poverty and adolescent adversity. Hence our hypothetical estimates should be regarded as upper bounds, designed to gauge the maximum possible reductions that could be achieved. Nor does our study identify and decipher the mechanisms and mediating pathways that link childhood household income and these adverse outcomes, as this is beyond the scope of this study. Recent studies have found that a substantial part of the relationship between socioeconomic conditions and adverse adolescent outcomes (about a fifth according to one recent study ^11^) can be explained (mediated) by adverse life experiences (ACEs), which include domestic violence and abuse, poor parental mental health, divorce and parental alcohol and substance use.^11 19 20^ The clustering of multiple adverse outcomes across adolescents experiencing early life poverty, which we find, could also be influenced by ACEs and clustering of multiple ACEs across adolescents, and it is of interest for future research to explore this further. Finally, our hypothetical levelling up scenarios are based on these simple associations and are un-costed. They are intended to provide upper bounds for policy makers interested in understanding the impacts that reducing childhood poverty and childhood income inequality might have on dealing with the burden of adverse adolescent outcomes rather than causal estimates of actual policies that could be implemented to achieve these ends.

### Implications

Our findings suggest that public health research needs to start looking more closely at the clustering of health risk factors, just as the study of multimorbidity has recently started to transform clinical research. Low household income in early childhood produces clustering of multiple adverse adolescent outcomes. Our modelling shows that simply raising the household income of our poorest families would not substantially change this observed pattern. The reduction of absolute poverty may be a necessary component of levelling up, but it is not sufficient. An ambitious levelling up agenda needs to recognise the complexities of childhood and tackle the multiple interacting and intersecting factors that determine the outcomes of children growing up in our most disadvantaged areas. Our findings provide evidence in support of whole system approaches to tackling multiple childhood inequalities (e.g. the Royal College of Paediatric and Child Health’s inequality programme) and coordinated multi-agency working between public services (e.g. health and education). No single agency is capable of addressing the complex problems impacting the early life experiences of children growing up in poverty. Current policy approaches implicitly assume that a single issue can be isolated and addressed by a given agency – for example, public health initiatives are largely disconnected from educational provision and vice versa. In more affluent households, this approach works because a childhood vulnerability is likely to occur in isolation and thus can be effectively mitigated through single agency responses. In disadvantaged communities, however, vulnerabilities multiply and traditional public service delivery models fail. This can help explain why relative poverty is often a better predictor of poor outcomes than absolute poverty. These conclusions are also consistent with recent calls for data driven place-based approaches in the UK’s most disadvantaged areas.^28^ These approaches could allow genuine multi-agency responses tailored to the context of specific communities and underpinned with the insights, evidence, and information sharing afforded through connected data.

### Declaration of interests

None of the authors have any conflict of interest to declare

### Data sharing

Data are available to researchers from the UK Data Service (beta.ukdataservice.ac.uk/datacatalogue/series/series?id=2000031)

## Data Availability

All data produced are available online

https://ukdataservice.ac.uk/

## Funding

UK Prevention Research Partnership (MR/S037527/1)

## Multiple imputations and weighting

As in all longitudinal studies there is attrition over time in the MCS, which if not addressed can bias results and reduce the statistical power of the sample. Of the 19,519 children who were initially recruited for the study only 10,757 cohort members provided any data at age 17, with even lower response rates on some specific survey questions. To deal with missing data, we used multiple imputation using chained equations to impute missing data back to the age 5 survey sample, which was the last data point for the income measure used in this study.^a^ ^b^ A number of auxiliary variables that were predictive of the five outcomes at age 17 were used in the imputation to improve the accuracy of our estimates.^c^ These variables were from various sweeps through the longitudinal study and included cognitive assessments, child mental health, physical health, substance use, and obesity. Using chained equations, 30 datasets were generated. Post-imputation, the final sample used in our analyses consisted of 15,245 cohort members. Weights included in the MCS dataset were used to adjust for attrition between the initial survey and the age 5 survey, and to adjust for the complex sampling design. Using these weights and the imputation strategy has allowed us to provide measures and estimate that are as close as possible to being nationally representative of this generation.

Notes:

**Table S1:** Main sample characteristics (N=15,245)

| **Sex** |  | % weighted |
| --- | --- | --- |
| Female | 7447 | 48.9% |
| Male | 7798 | 51.1% |
| **Ethnicity** |  |  |
| White | 12706 | 86.7% |
| Mixed | 443 | 3.2% |
| Indian | 382 | 1.8% |
| Pakistani and Bangladeshi | 987 | 4.2% |
| Black and Black British | 514 | 2.8% |
| Other ethnic group | 213 | 1.2% |
| **Country** |  |  |
| England | 9716 | 82.3% |
| Wales | 2181 | 4.8% |
| Scotland | 1814 | 9.0% |
| Northern Ireland | 1534 | 3.8% |
| **Childhood household income quintile group** |  |  |
| Lowest 20% [£117 weekly average] [range: £0-154] | 3070 | 17.6% |
| Lowest 20-40% [£191 weekly average] [range: £155-233] | 3093 | 18.7% |
| Middle 40-60% [£285 weekly average] [range: £234-337] | 3068 | 20.4% |
| Highest 60-80% [£398 weekly average] [range: £338-470] | 3001 | 20.8% |
| Highest 80-100% [£644 weekly average][range: £471-1195] | 3013 | 22.5% |
| **Individual adverse adolescent outcomes at age 17** |  |  |
| Psychological distress | 2222 | 14.7% |
| Self-reported poor health | 1144 | 7.6% |
| Regular smoker | 1529 | 10.4% |
| Obese | 3717 | 23.3% |
| Poor academic achievement | 5785 | 37.0% |
| **Index of adverse adolescent outcomes at age 17** |  |  |
| Zero | 6046 | 40.8% |
| One | 5428 | 34.9% |
| Two | 2625 | 16.9% |
| Three | 1145 | 5.8% |
| Four | 217 | 1.4% |
| Five | 33 | 0.2% |

Note: The weighted percentages are adjusted for the complex sampling design and for attrition between the initial survey and the age 5 survey used as the imputation sample.

**Table S2:** Crosstabulation of early childhood income (age 0-5) and the five adverse outcomes at age 17.

**Panel A**
| **Whole sample (15,245)** | **PSYCHOLOGICAL DISTRESS** |  |  |
| --- | --- | --- | --- |
|  | No | Yes | Total |
| **Income quintiles** |  |  |  |
| **Lowest 20%** | 2223 | 455 | 2678 |
| Row (within income group) | 83.0% [81.0-85.1] | 17.0% [14.9-19.0] | 100.0% |
| Col (within response) | 17.1% [15.6-18.6] | 20.3% [17.6-23.1] | 17.6% |
| **Lowest 20-40%** | 2384 | 472 | 2855 |
| Row (within income group) | 83.5% [81.4-85.5] | 16.5% [14.5-18.6] | 100.0% |
| Col (within response) | 18.3% [17.2-19.5] | 21.1% [18.5-23.6] | 18.7% |
| **Middle 40-60%** | 2629 | 484 | 3114 |
| Row (within income group) | 84.4% [82.6-86.3] | 15.6% [13.7-17.4] | 100.0% |
| Col (within response) | 20.2% [19.0-21.4] | 21.7% [19.0-24.3] | 20.4% |
| **Highest 60-80%** | 2738 | 427 | 3165 |
| Row (within income group) | 86.5% [84.9-88.1] | 13.5% [11.9-15.1] | 100.0% |
| Col (within response) | 21.0% [21.9-22.2] | 19.1% [16.7-21.4] | 20.8% |
| **Highest 80-100%** | 3034.8 | 398.8 | 3433.6 |
| Row (within income group) | 88.4% [87.1-89.7] | 11.6% [10.3-12.9] | 100.0% |
| Col (within response) | 23.3% [21.1-25.6] | 17.8% [15.0-20.6] | 22.5% |
| **Total** | 13009 | 2236 | 15245 |
|  | 85.3% [84.6-86.1] | 14.7% [13.9-15.4] | 100.0% |
|  | 100.0% | 100.0% |  |

(Table S2 continued) **Panel B**
| Whole sample (15,245) | POOR HEALTH |  |  |
| --- | --- | --- | --- |
|  | No | Yes | Total |
| **Income quintiles** |  |  |  |
| **Lowest 20%** | 2353 | 324 | 2678 |
| Row (within income group) | 87.9% [85.9-89.9] | 12.1% [10.1-14.1] | 100.0% |
| Col (within response) | 16.7% [15.2-18.2] | 27.8% [23.6-32.1] | 17.6% |
| **Lowest 20-40%** | 2579 | 276 | 2855 |
| Row (within income group) | 90.3% [88.6-92.0] | 9.7% [8.0-11.4] | 100.0% |
| Col (within response) | 18.3% [17.2-19.5] | 23.7% [20.3-27.1] | 18.7% |
| **Middle 40-60%** | 2893 | 221 | 3114 |
| Row (within income group) | 92.9% [91.5-94.3] | 7.1% [5.7-8.5] | 100.0% |
| Col (within response) | 20.5% [19.3-21.8] | 18.9% [15.2-22.5] | 20.4% |
| **Highest 60-80%** | 2975 | 190 | 3165 |
| Row (within income group) | 94.0% [92.8-95.2] | 6.0% [4.8-7.2] | 100.0% |
| Col (within response) | 21.1% [20.0-22.3] | 16.3% [13.3-19.3] | 20.8% |
| **Highest 80-100%** | 3280 | 154 | 3434 |
| Row (within income group) | 95.5% [94.6-96.5] | 4.5% [3.5-5.4] | 100.0% |
| Col (within response) | 23.3% [21.0-25.5] | 13.2% [10.2-16.2] | 22.5% |
| **Total** | 14079 | 1166 | 15245 |
|  | 92.4% [91.6-93.1] | 7.6% [6.9-8.4] | 100.0% |
|  | 100.0% | 100.0% |  |

(Table S2 continued) **Panel C**
| **Whole sample (15,245)** | **OBESITY** |  |  |
| --- | --- | --- | --- |
| **Income quintiles** | No | Yes | Total |
| **Lowest 20%** | 1890 | 788 | 2678 |
| Row (within income group) | 70.6% [68.1-73.1] | 29.4% [26.9-31.9] | 100.0% |
| Col (within response) | 16.2% [14.7-17.6] | 22.2% [19.9-24.5] | 17.6% |
| **Lowest 20-40%** | 2035 | 820 | 2855 |
| Row (within income group) | 71.3% [69.0-73.6] | 28.7% [26.4-31.0] | 100.0% |
| Col (within response) | 17.4% [16.2-18.6] | 23.1% [21.2-25.0] | 18.7% |
| **Middle 40-60%** | 2351 | 762 | 3114 |
| Row (within income group) | 75.5% [73.5-77.6] | 24.5% [22.4-26.5] | 100.0% |
| Col (within response) | 20.1% [18.8-21.4] | 21.5% [19.7-23.3] | 20.4% |
| **Highest 60-80%** | 2504 | 661 | 3165 |
| Row (within income group) | 79.1% [77.3-80.9] | 20.9% [19.1-22.7] | 100.0% |
| Col (within response) | 21.4% [20.2-22.6] | 18.6% [16.8-20.5] | 20.8% |
| **Highest 80-100%** | 2915 | 518 | 3434 |
| Row (within income group) | 84.9% [83.2-86.6] | 15.1% [13.4-16.8] | 100.0% |
| Col (within response) | 24.9% [22.4-27.4] | 14.6% [12.7-16.6] | 22.5% |
| **Total** | 11696 | 3549 | 15245 |
|  | 76.7% [75.6-77.8] | 23.3% [22.2-24.4] | 100.0% |
|  | 100.0% | 100.0% |  |

(Table S2 continued) **Panel D**
| **Whole sample (15,245)** | **SMOKING** |  |  |
| --- | --- | --- | --- |
| **Income quintiles** | No | Yes | Total |
| **Lowest 20%** | 2182 | 495 | 2678 |
| Row (within income group) | 81.5% [78.9-84.1] | 18.5% [15.9-21.1] | 100.0% |
| Col (within response) | 16.0% [14.4-17.5] | 31.2% [27.5-35.0] | 17.6% |
| **Lowest 20-40%** | 2448 | 408 | 2855 |
| Row (within income group) | 85.7% [83.8-87.7] | 14.3% [12.2-16.2] | 100.0% |
| Col (within response) | 17.9% [16.8-19.1] | 25.7% [22.7-28.7] | 18.7% |
| **Middle 40-60%** | 2806 | 307 | 3114 |
| Row (within income group) | 90.1% [88.6-91.6] | 9.9% [8.4-11.4] | 100.0% |
| Col (within response) | 20.5% [19.3-21.8] | 19.4% [16.6-22.2] | 20.4% |
| **Highest 60-80%** | 2969 | 196 | 3165 |
| Row (within income group) | 93.8% [92.5-95.1] | 6.2% [4.9-7.5] | 100.0% |
| Col (within response) | 21.7% [20.5-22.9] | 12.4% [10.1-14.7] | 20.8% |
| **Highest 80-100%** | 3255 | 178 | 3434 |
| Row (within income group) | 94.8% [93.7-95.9] | 5.2% [4.1-6.3] | 100.0% |
| Col (within response) | 23.8% [21.6-26.1] | 11.3% [8.6-13.9] | 22.5% |
| **Total** | 13660 | 1585 | 15245 |
|  | 89.6% [88.8-90.5] | 10.4% [9.5-11.2] | 100.0% |
|  | 100.0% | 100.0% |  |

(Table S2 continued) **Panel E**
| **Whole sample (15,245)** | **POOR ACADEMIC ACHIEVEMENT** |  |  |
| --- | --- | --- | --- |
|  | No | Yes | Total |
| **Income quintiles** |  |  |  |
| **Lowest 20%** | 976 | 1702 | 2678 |
| Row (within income group) | 36.4% [33.7-39.2] | 63.6% [60.8-66.3] | 100.0% |
| Col (within response) | 10.2% [8.8-11.5] | 30.2% [28.0-32.4] | 17.6% |
| **Lowest 20-40%** | 1376 | 1479 | 2855 |
| Row (within income group) | 48.2% [45.5-50.9] | 51.8% [49.1-54.5] | 100.0% |
| Col (within response) | 14.3% [13.2-15.5] | 26.2% [24.6-27.9] | 18.7% |
| **Middle 40-60%** | 1964 | 1150 | 3114 |
| Row (within income group) | 63.1% [60.7-65.5] | 36.9% [34.5-39.3] | 100.0% |
| Col (within response) | 20.4% [18.9-21.9] | 20.4% [19.0-21.8] | 20.4% |
| **Highest 60-80%** | 2338 | 827 | 3165 |
| Row (within income group) | 73.9% [71.7-76.1] | 26.1% [23.9-28.3] | 100.0% |
| Col (within response) | 24.3% [22.9-25.8] | 14.7% [13.3-16.1] | 20.8% |
| **Highest 80-100%** | 2955 | 478 | 3434 |
| Row (within income group) | 86.1% [84.2-87.9] | 13.9% [12.1-15.8] | 100.0% |
| Col (within response) | 30.8% [28.0-33.5] | 8.5% [7.3-9.7] | 22.5% |
| **Total** | 9609 | 5636 | 15245 |
|  | 63.0% [61.3-64.7] | 37.0% [35.3-38.7] | 100.0% |
|  | 100.0% | 100.0% |  |

Notes: weighted for attrition and sampling design

**Table S3:**
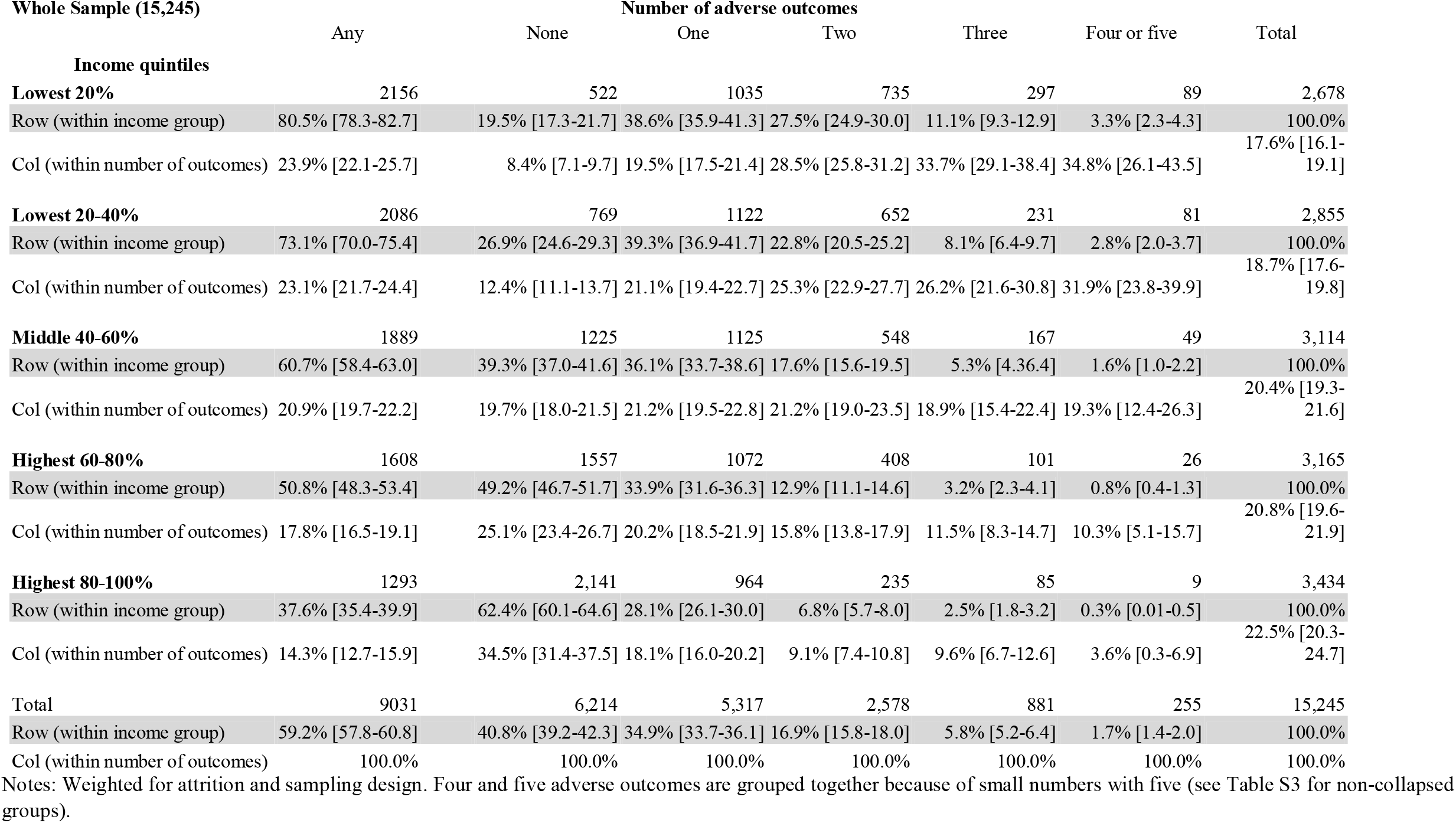
Crosstabulation of early childhood income (age 0-5) and number of adverse outcomes at age 17.

**Table S4:**
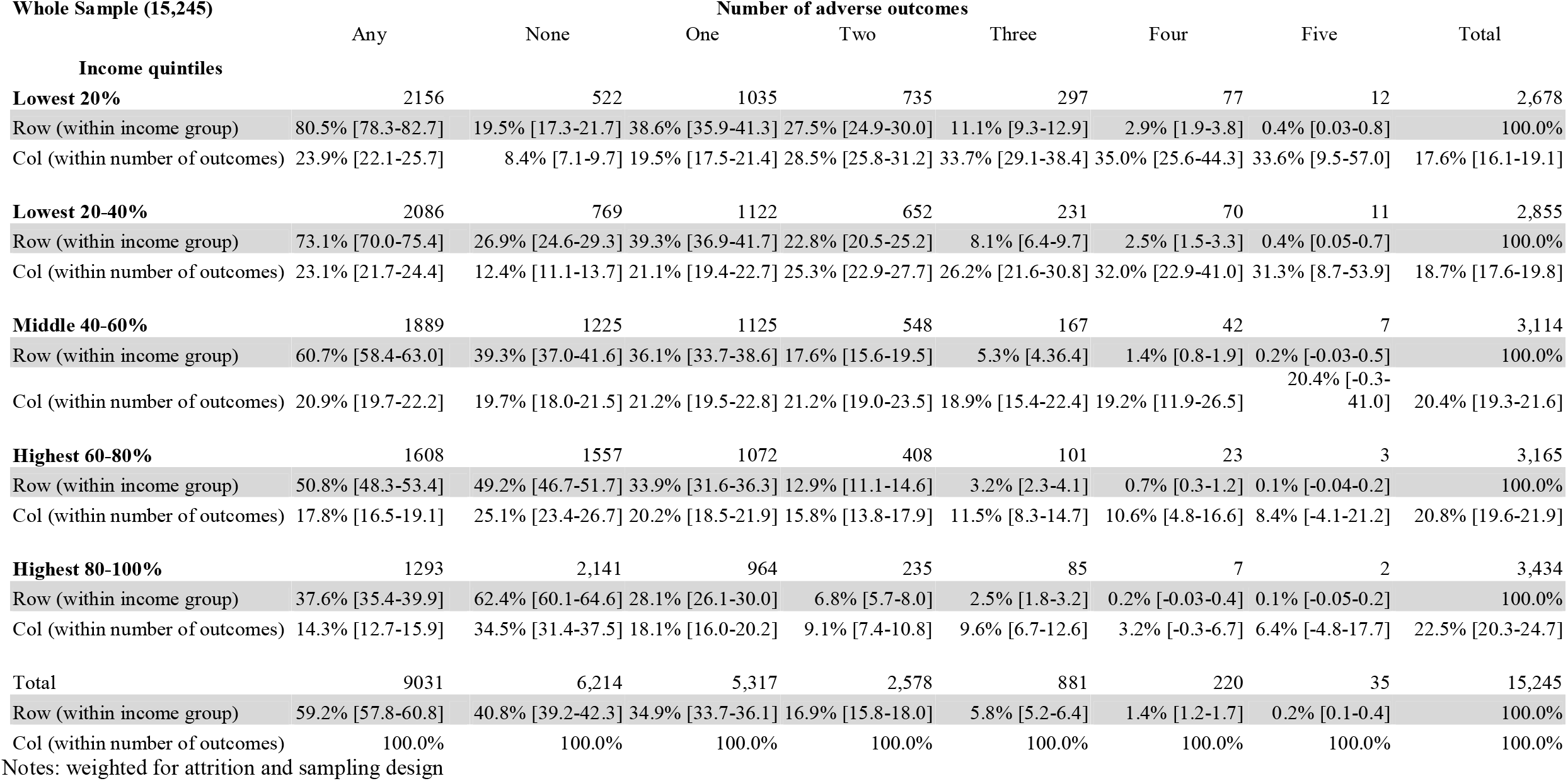
Crosstabulation of early childhood income (age 0-5) and number of adverse outcomes at age 17 (without collapsing four and five adversities)

**Table S5:**
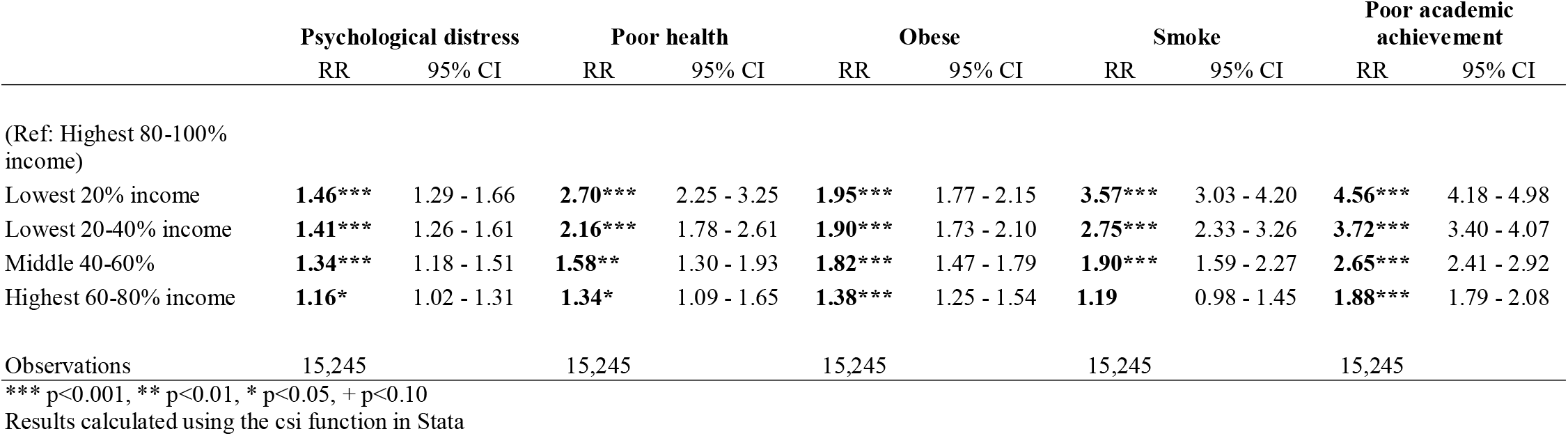
Risk ratios with confidence intervals of income inequalities in each adverse outcome at age 17.

**Table S6:** Risk ratios with confidence intervals of income inequalities in number of adverse outcomes at age 17.

|  | Any |  | None |  | One |  | Two |  | Three |  | Four or five |  |
| --- | --- | --- | --- | --- | --- | --- | --- | --- | --- | --- | --- | --- |
|  | RR | 95% CI | RR | 95% CI | RR | 95% CI | RR | 95% CI | RR | 95% CI | RR | 95% CI |
| (Ref: Highest 80-100% income) |  |  |  |  |  |  |  |  |  |  |  |  |
| Lowest 20% income | **2.14\*\*\*** | 2.04 - 2.24 | **0.31\*\*\*** | 0.29 - 0.34 | **1.38\*\*\*** | 1.28 - 1.50 | **4.01\*\*\*** | 3.50 - 4.61 | **4.48\*\*\*** | 3.54 - 5.67 | **12.68\*\*\*** | 6.40 - 25.12 |
| Lowest 20-40% income | **1.94\*\*\*** | 1.85 - 2.04 | **0.43\*\*\*** | 0.41 - 0.46 | **1.40\*\*\*** | 1.30 - 1.50 | **3.34\*\*\*** | 2.90 - 3.84 | **3.27\*\*\*** | 2.56 - 4.17 | **10.82\*\*\*** | 5.45 - 21.51 |
| Middle 40-60% | **1.61\*\*\*** | 1.53 - 1.70 | **0.63\*\*\*** | 0.60 - 0.66 | **1.29\*\*\*** | 1.20 - 1.38 | **2.57\*\*\*** | 2.22 - 2.97 | **2.17\*\*\*** | 1.67 - 2.80 | **6.00\*\*\*** | 2.95 - 12.20 |
| Highest 60-80% income | **1.35\*\*\*** | 1.28 - 1.43 | **0.79\*\*\*** | 0.76 - 0.82 | **1.21\*\*\*** | 1.12 - 1.30 | **1.88\*\*\*** | 1.62 - 2.20 | 1.29 | 0.97 - 1.71 | **3.13\*** | 1.47 - 6.68 |
| Observations | 15,245 |  | 15,245 |  | 15,245 |  | 15,245 |  | 15,245 |  | 15,245 |  |

\*\*\* p<0.001, \*\* p<0.01, \* p<0.05, + p<0.10
Results calculated using the csi function in Stata

**Figure S1:**
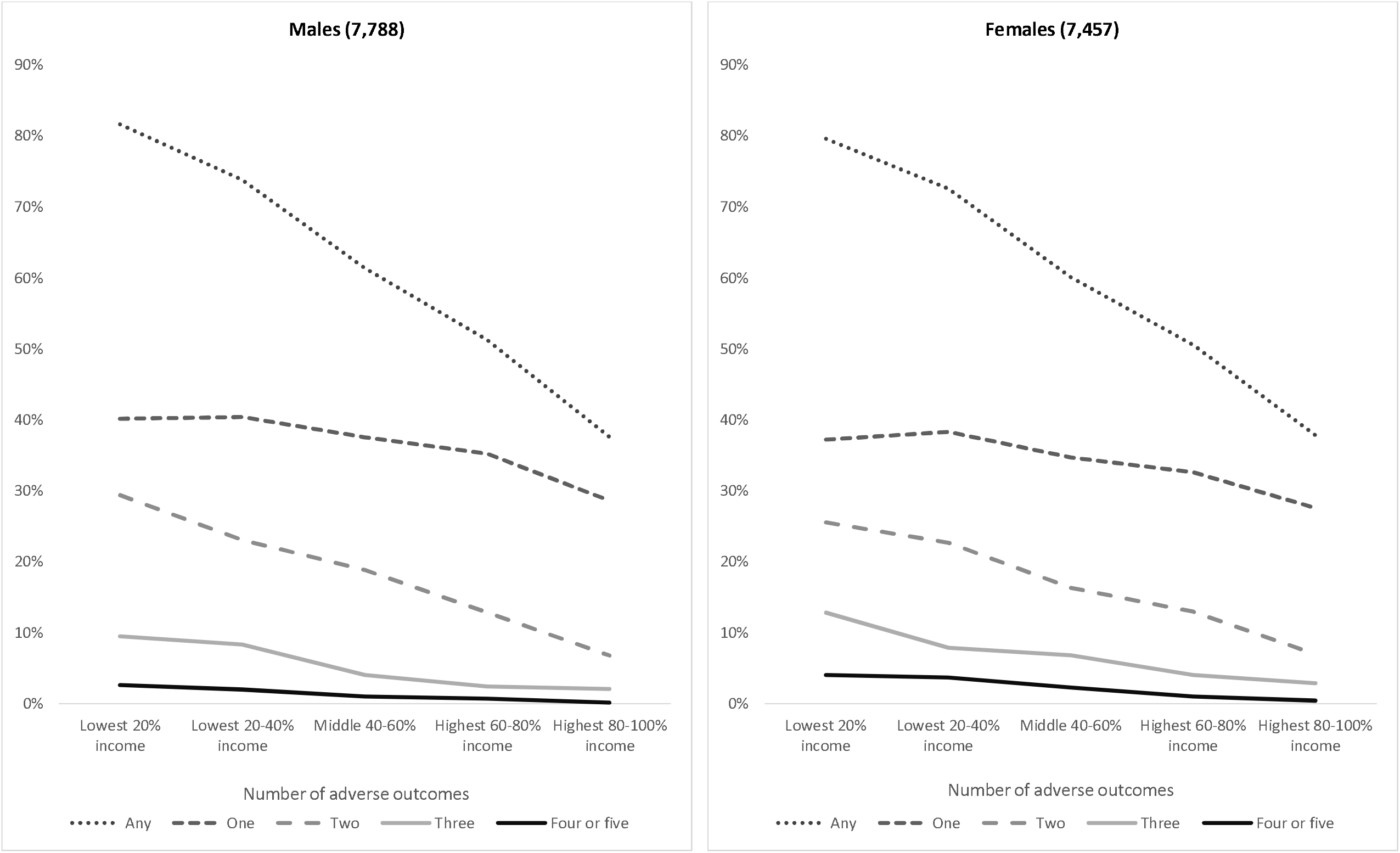
Absolute differences – proportion with number of adverse outcomes at age 17 in each income group - by gender.

**Figure S2:**
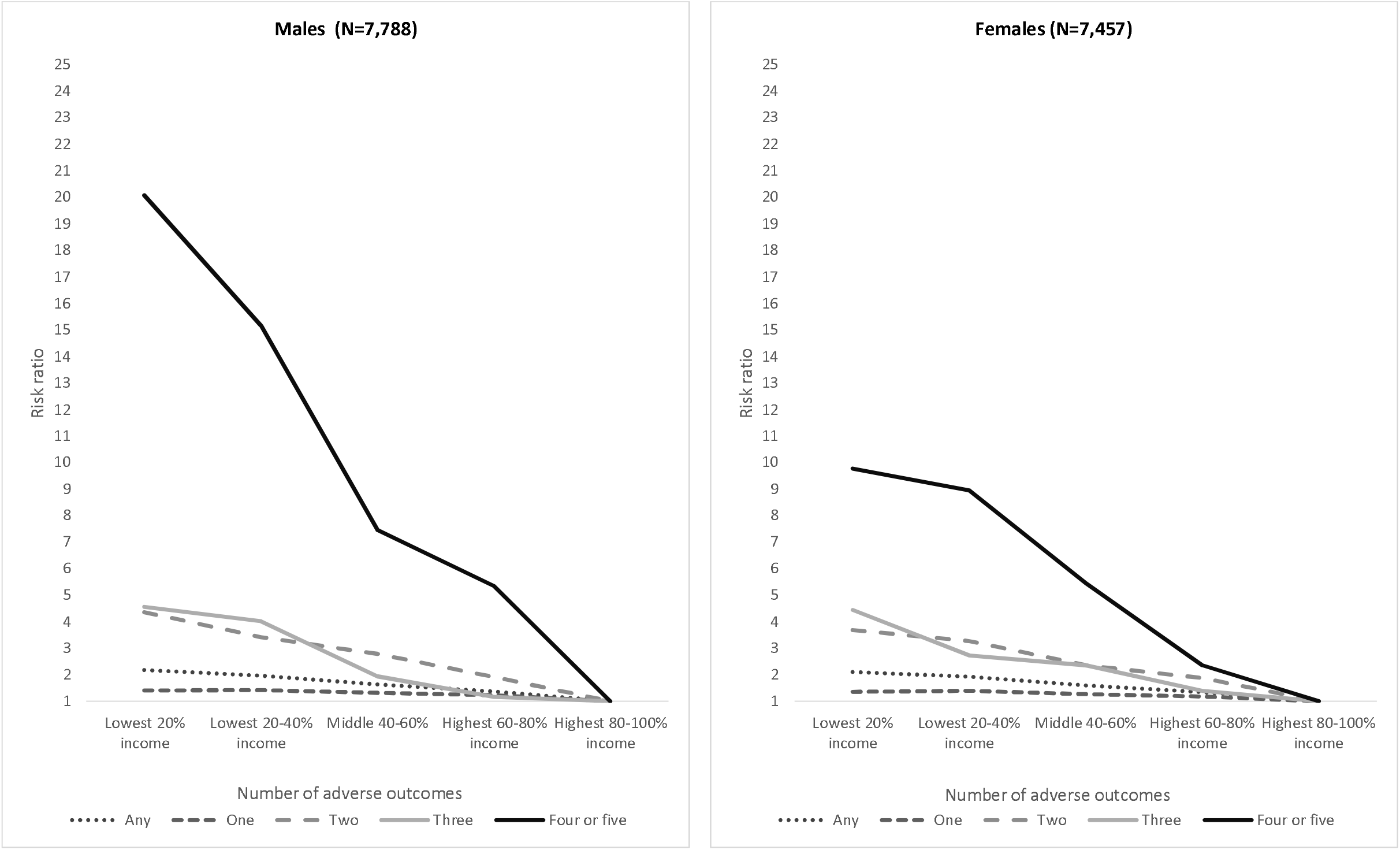
Relative differences - risk ratio differences in number of adverse outcomes at age 17 between income groups (ref. highest quintile) - by gender.

**Table S7:**
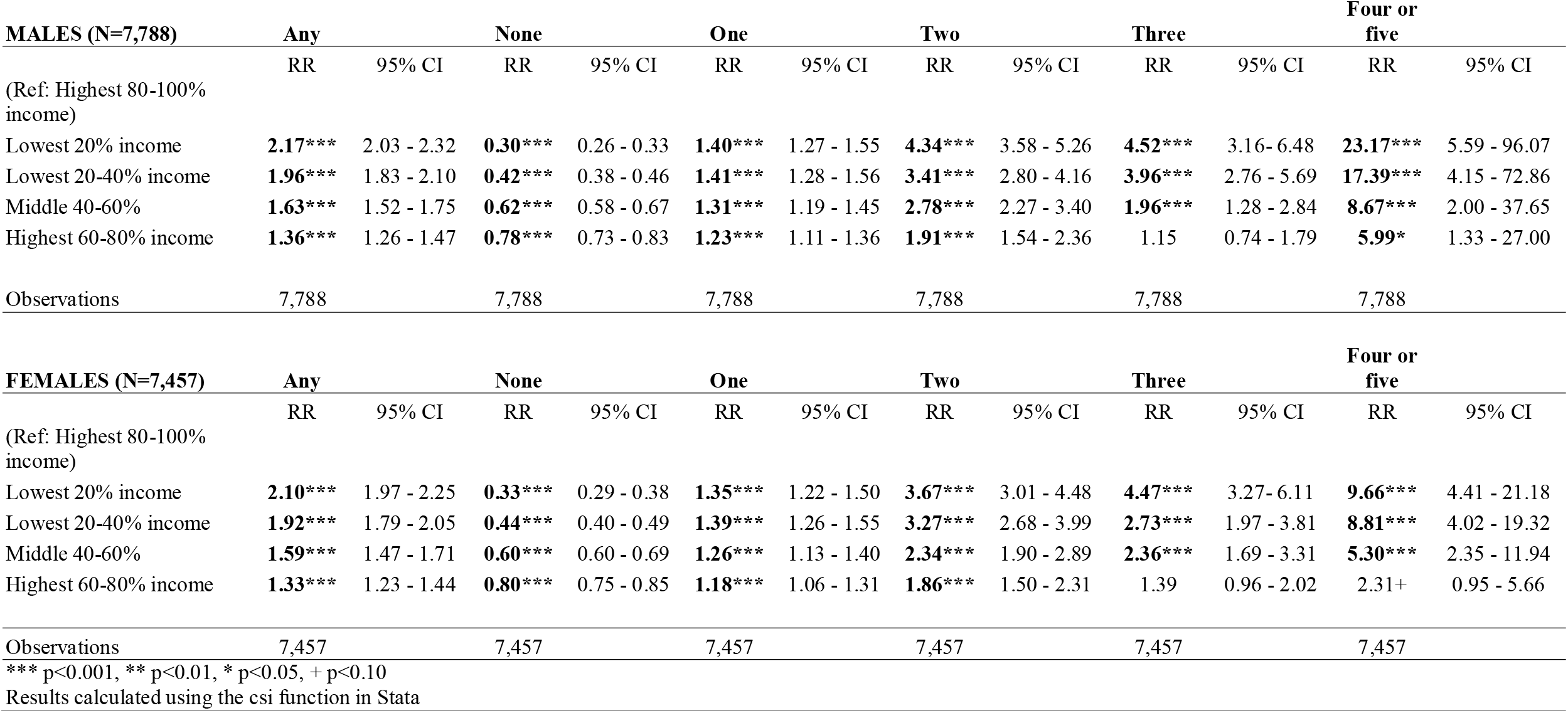
Risk ratios with confidence intervals of income inequalities in adverse outcomes at age 17 (by gender)

| **MALES (N=7,788)** | **Any** |  | **None** |  | **One** |  | **Two** |  | **Three** |  | **Four or five** |  |
| --- | --- | --- | --- | --- | --- | --- | --- | --- | --- | --- | --- | --- |
|  | RR | 95% CI | RR | 95% CI | RR | 95% CI | RR | 95% CI | RR | 95% CI | RR | 95% CI |
| (Ref: Highest 80-100% income) |  |  |  |  |  |  |  |  |  |  |  |  |
| Lowest 20% income | **2.17\*\*\*** | 2.03 - 2.32 | **0.30\*\*\*** | 0.26 - 0.33 | **1.40\*\*\*** | 1.27 - 1.55 | **4.34\*\*\*** | 3.58 - 5.26 | **4.52\*\*\*** | 3.16 - 6.48 | **23.17\*\*\*** | 5.59 - 96.07 |
| Lowest 20-40% income | **1.96\*\*\*** | 1.83 - 2.10 | **0.42\*\*\*** | 0.38 - 0.46 | **1.41\*\*\*** | 1.28 - 1.56 | **3.41\*\*\*** | 2.80 - 4.16 | **3.96\*\*\*** | 2.76 - 5.69 | **17.39\*\*\*** | 4.15 - 72.86 |
| Middle 40-60% | **1.63\*\*\*** | 1.52 - 1.75 | **0.62\*\*\*** | 0.58 - 0.67 | **1.31\*\*\*** | 1.19 - 1.45 | **2.78\*\*\*** | 2.27 - 3.40 | **1.96\*\*\*** | 1.28 - 2.84 | **8.67\*\*\*** | 2.00 - 37.65 |
| Highest 60-80% income | **1.36\*\*\*** | 1.26 - 1.47 | **0.78\*\*\*** | 0.73 - 0.83 | **1.23\*\*\*** | 1.11 - 1.36 | **1.91\*\*\*** | 1.54 - 2.36 | 1.15 | 0.74 - 1.79 | **5.99\*** | 1.33 - 27.00 |
| Observations | 7,788 |  | 7,788 |  | 7,788 |  | 7,788 |  | 7,788 |  | 7,788 |  |

| **FEMALES (N=7,457)** | **Any** |  | **None** |  | **One** |  | **Two** |  | **Three** |  | **Four or five** |  |
| --- | --- | --- | --- | --- | --- | --- | --- | --- | --- | --- | --- | --- |
|  | RR | 95% CI | RR | 95% CI | RR | 95% CI | RR | 95% CI | RR | 95% CI | RR | 95% CI |
| (Ref: Highest 80-100% income) |  |  |  |  |  |  |  |  |  |  |  |  |
| Lowest 20% income | **2.10\*\*\*** | 1.97 - 2.25 | **0.33\*\*\*** | 0.29 - 0.38 | **1.35\*\*\*** | 1.22 - 1.50 | **3.67\*\*\*** | 3.01 - 4.48 | **4.47\*\*\*** | 3.27 - 6.11 | **9.66\*\*\*** | 4.41 - 21.18 |
| Lowest 20-40% income | **1.92\*\*\*** | 1.79 - 2.05 | **0.44\*\*\*** | 0.40 - 0.49 | **1.39\*\*\*** | 1.26 - 1.55 | **3.27\*\*\*** | 2.68 - 3.99 | **2.73\*\*\*** | 1.97 - 3.81 | **8.81\*\*\*** | 4.02 - 19.32 |
| Middle 40-60% | **1.59\*\*\*** | 1.47 - 1.71 | **0.60\*\*\*** | 0.60 - 0.69 | **1.26\*\*\*** | 1.13 - 1.40 | **2.34\*\*\*** | 1.90 - 2.89 | **2.36\*\*\*** | 1.69 - 3.31 | **5.30\*\*\*** | 2.35 - 11.94 |
| Highest 60-80% income | **1.33\*\*\*** | 1.23 - 1.44 | **0.80\*\*\*** | 0.75 - 0.85 | **1.18\*\*\*** | 1.06 - 1.31 | **1.86\*\*\*** | 1.50 - 2.31 | 1.39 | 0.96 - 2.02 | 2.31+ | 0.95 - 5.66 |
| Observations | 7,457 |  | 7,457 |  | 7,457 |  | 7,457 |  | 7,457 |  | 7,457 |  |

\*\*\* p<0.001, \*\* p<0.01, \* p<0.05, + p<0.10 Results calculated using the csi function in Stata

**Figure S3:**
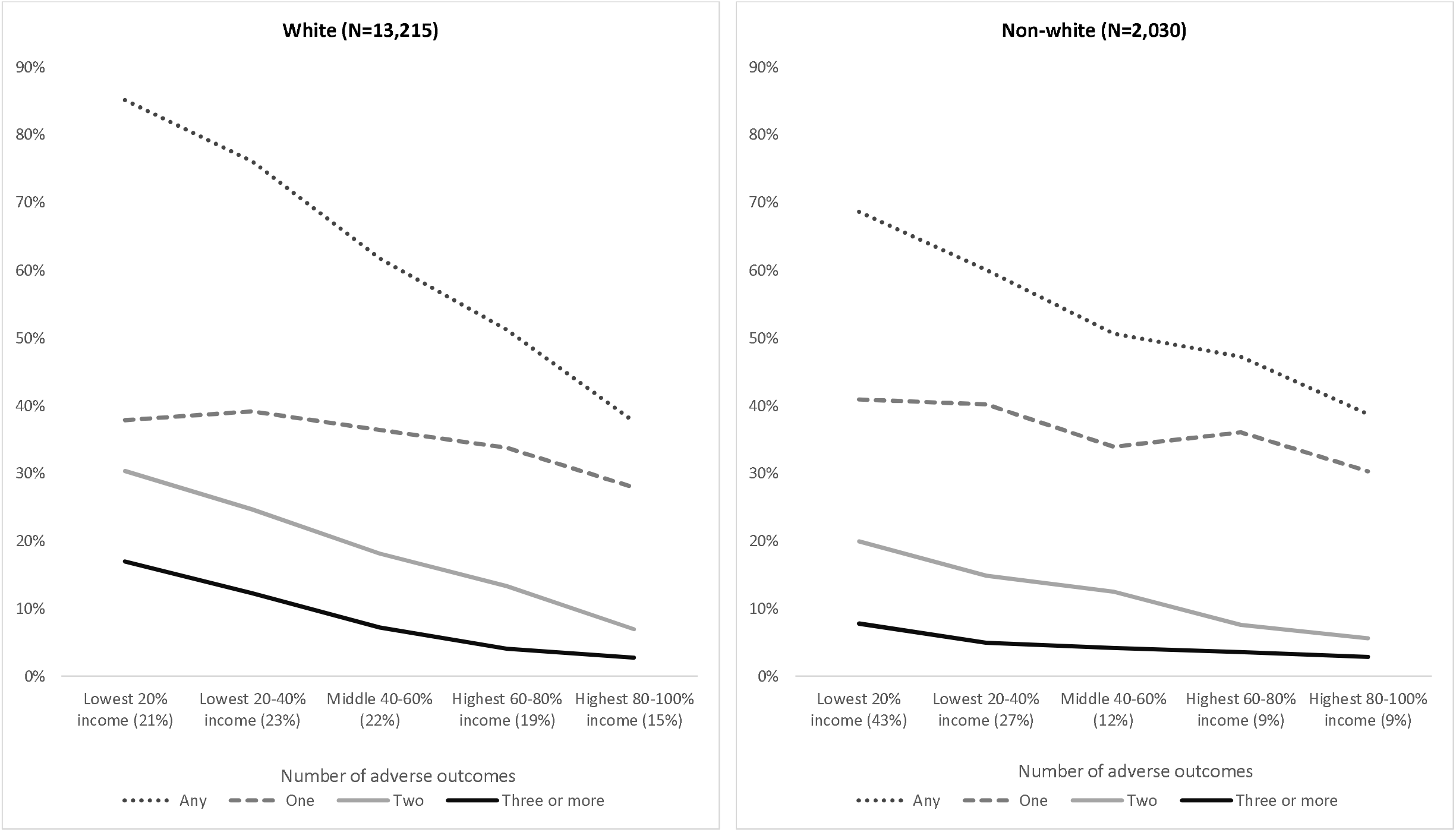
Absolute differences – proportion with number of adverse outcomes at age 17 in each income group - by ethnicity. Note: Percentages in parenthesis for each income quintile show the income distribution for white and non-white. We see how non-white ethnic groups are overrepresented (43%) in the lowest income quintile.

**Figure S4:**
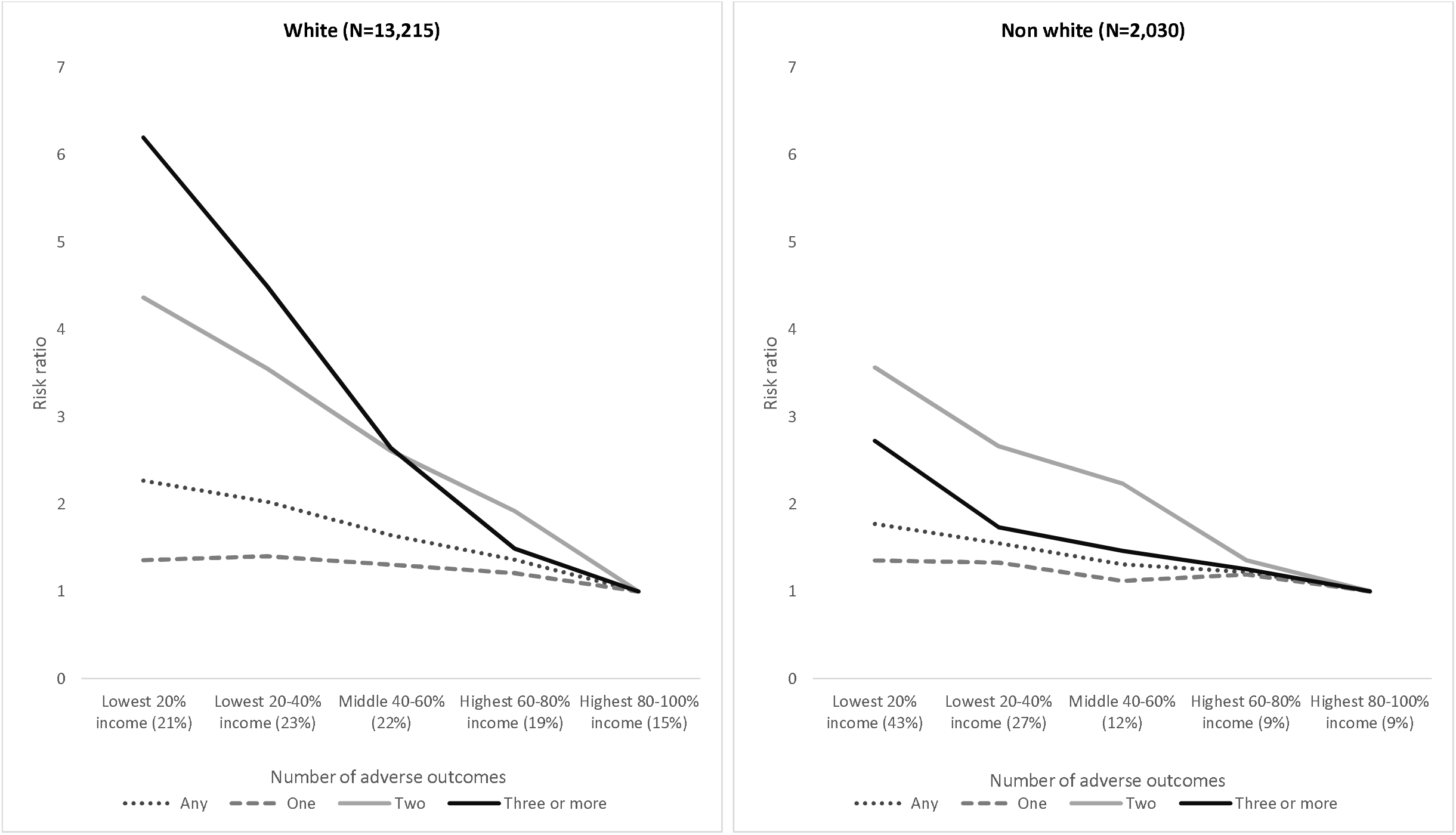
Relative differences - risk ratio differences in number of adverse outcomes at age 17 between income groups (ref. highest quintile) - by ethnicity. Note: Percentages in parenthesis for each income quintile show the income distribution for white and non-white. We see how non-white ethnic groups are overrepresented (43%) in the lowest income quintile.

**Table S8:** Risk ratios with confidence intervals of income inequalities in adverse outcomes at age 17 (by ethnicity)

| **WHITE (N=13,215)** | **Any** |  | **None** |  | **One** |  | **Two** |  | **Three or more** |  |
| --- | --- | --- | --- | --- | --- | --- | --- | --- | --- | --- |
|  | RR | 95% CI | RR | 95% CI | RR | 95% CI | RR | 95% CI | RR | 95% CI |
| (Ref: Highest 80-100% income) |  |  |  |  |  |  |  |  |  |  |
| Lowest 20% income | **2.26\*\*\*** | 2.16 - 2.38 | **0.24\*\*\*** | 0.22 - 0.27 | **1.36\*\*\*** | 1.25 - 1.47 | **4.37\*\*\*** | 3.78 - 5.04 | **6.17\*\*\*** | 4.91 - 7.76 |
| Lowest 20-40% income | **2.02\*\*\*** | 1.92 - 2.13 | **0.38\*\*\*** | 0.36 - 0.42 | **1.40\*\*\*** | 1.30 - 1.51 | **3.55\*\*\*** | 3.07 - 4.11 | **4.47\*\*\*** | 3.54 - 5.65 |
| Middle 40-60% | **1.64\*\*\*** | 1.56 - 1.73 | **0.61\*\*\*** | 0.58 - 0.65 | **1.30\*\*\*** | 1.21 - 1.40 | **2.61\*\*\*** | 2.25 - 3.03 | **2.63\*\*\*** | 2.06 - 3.36 |
| Highest 60-80% income | **1.36\*\*\*** | 1.29 - 1.44 | **0.78\*\*\*** | 0.75 - 0.82 | **1.21\*\*\*** | 1.12 - 1.30 | **1.92\*\*\*** | 1.64 - 2.25 | **1.48\*\*** | 1.13 - 1.94 |
| Observations | 13,215 |  | 13,215 |  | 13,215 |  | 13,215 |  | 13,215 |  |

| **NON-WHITE (N=2,030)** | **Any** |  | **None** |  | **One** |  | **Two** |  | **Three or more** |  |
| --- | --- | --- | --- | --- | --- | --- | --- | --- | --- | --- |
|  | RR | 95% CI | RR | 95% CI | RR | 95% CI | RR | 95% CI | RR | 95% CI |
| (Ref: Highest 80-100% income) |  |  |  |  |  |  |  |  |  |  |
| Lowest 20% income | **1.78\*\*\*** | 1.51 - 2.09 | **0.51\*\*\*** | 0.44 - 0.59 | **1.36\*\*** | 1.11 - 1.66 | **3.47\*\*\*** | 2.08 - 5.80 | **2.92\*\*** | 1.35 - 6.33 |
| Lowest 20-40% income | **1.56\*\*\*** | 1.31 - 1.84 | **0.65\*\*\*** | 0.57 - 0.75 | **1.33\*\*** | 1.08 - 1.65 | **2.61\*\*\*** | 1.53 - 4.44 | 1.86 | 0.82 - 4.24 |
| Middle 40-60% | **1.31\*\*** | 1.08 - 1.59 | **0.81\*\*** | 0.69 - 0.94 | 1.12 | 0.88 - 1.43 | **2.21\*\*** | 1.24 - 3.93 | 1.58 | 0.63 - 3.94 |
| Highest 60-80% income | 1.22+ | 0.99 - 1.50 | 0.86 | 0.74 - 1.00 | 1.20 | 0.93 - 1.54 | 1.35 | 0.70 - 2.62 | 1.29 | 0.48 - 3.50 |
| Observations | 2,030 |  | 2,030 |  | 2,030 |  | 2,030 |  | 2,030 |  |

\*\*\* p<0.001, \*\* p<0.01, \* p<0.05, + p<0.10 Results calculated using the csi function in Stata

**Figure S5:**
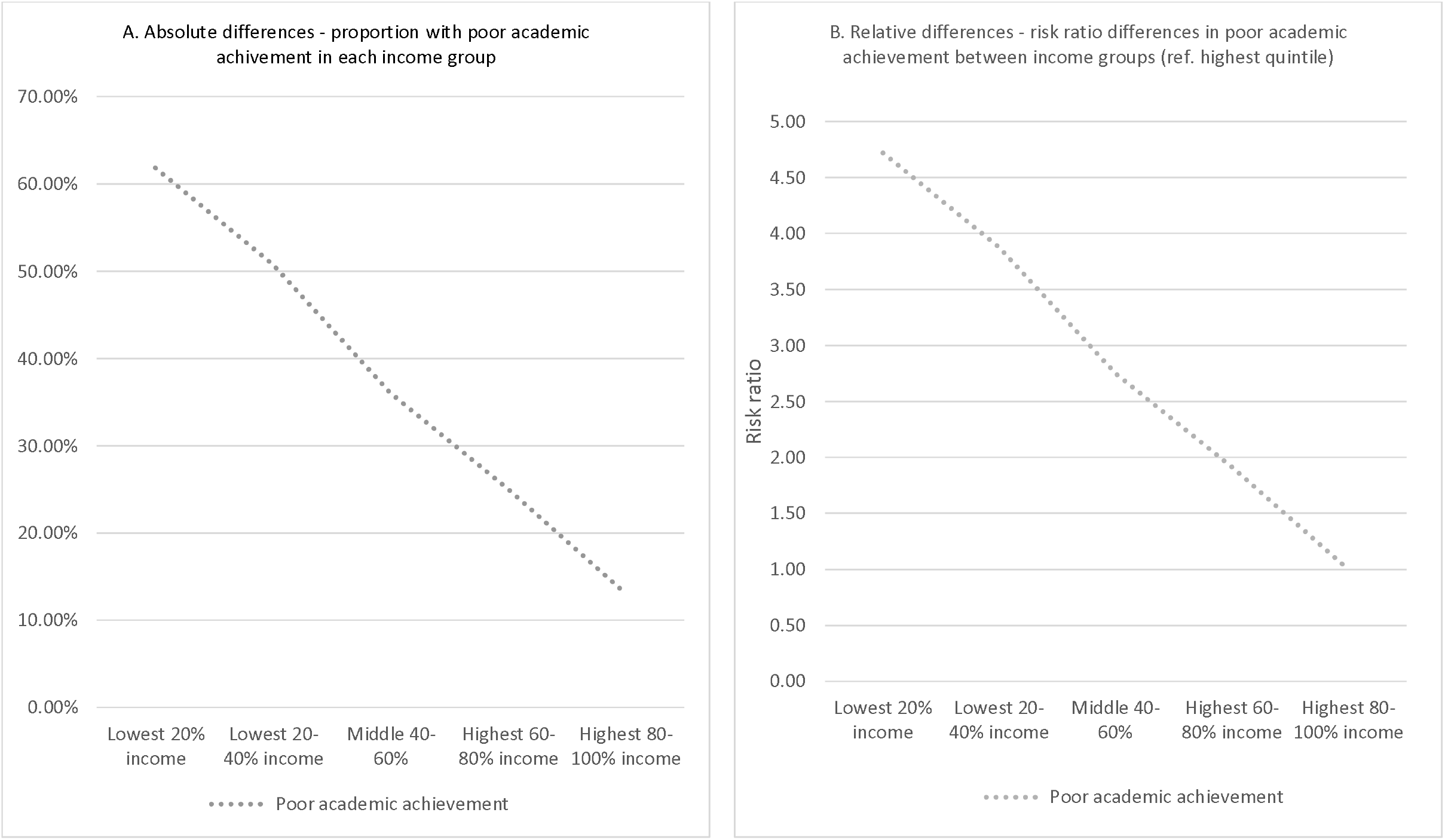
Poor academic achievement at age 17 by early childhood income (England, Wales and Northern Ireland, N=13,441)

**Figure S6:**
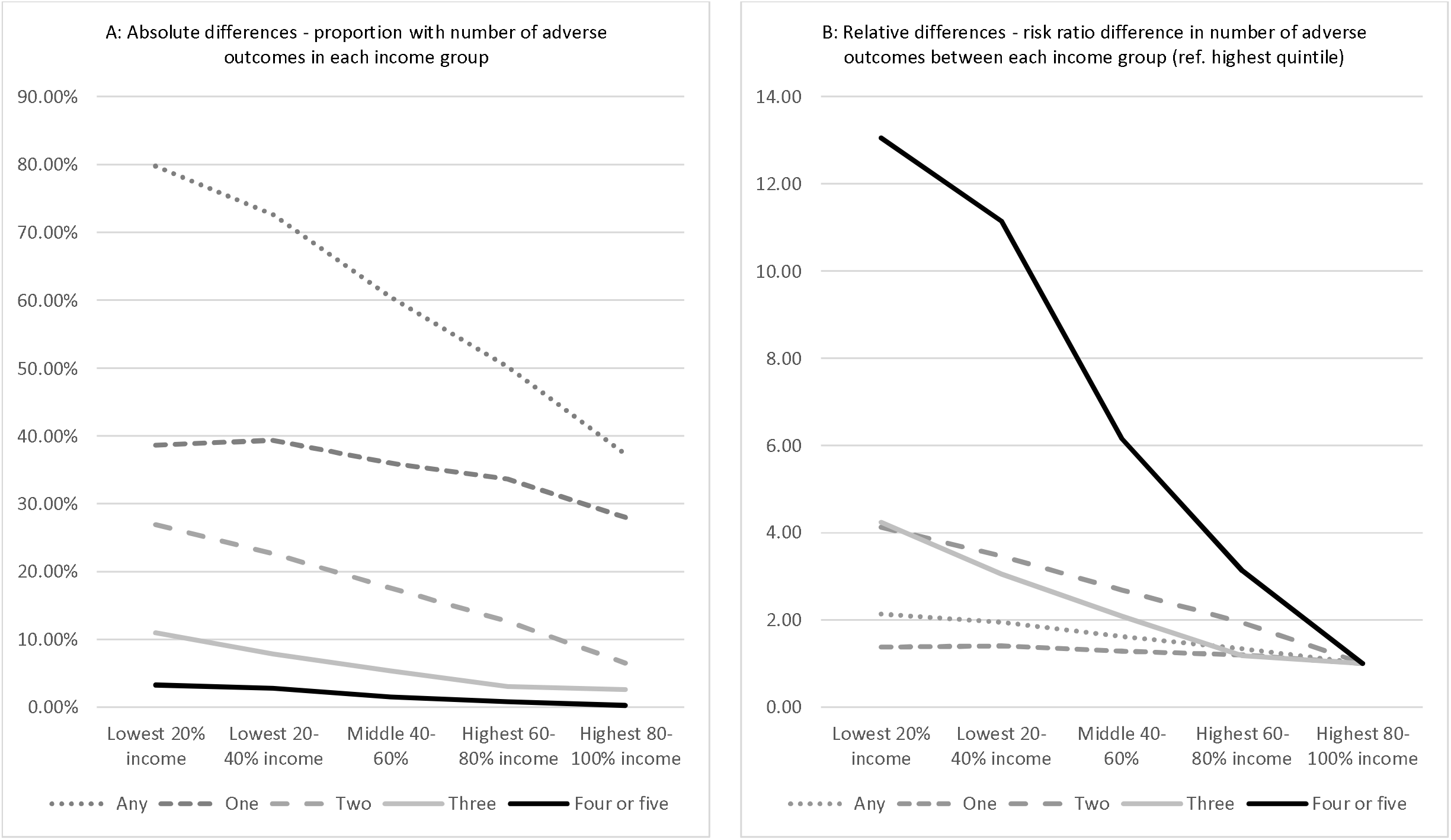
Number of adverse outcomes at age 17 by early childhood income (England, Wales and Northern Ireland, N=13,441)

**Table S9:** Results of levelling up scenarios – reduction in adverse health outcomes at age 17.

| **Panel A Proportional reduction in adverse outcomes from three hypothetical levelling up scenarios** |  |  |  |  |  |
| --- | --- | --- | --- | --- | --- |
|  | Psychological distress | Poor health | Obese | Smoke | Poor academic achievement |
| Scenario 1 | 0.55% [-6.44% -- 7.55%] | 5.60% [-6.50% -- 17.69%] | 0.54% [-4.57% -- 5.65%] | 7.13% [-4.01% -- 18.26%] | 5.58% [1.85% -- 9.30%] |
| Scenario 2 | 2.93% [-6.88% -- 12.74%] | 17.88% [1.96% -- 33.80%] | 7.14% [0.15% -- 14.14%] | 22.51% [8.43% -- 36.59%] | 20.19% [16.48% -- 23.89%] |
| Scenario 3 | 20.81% [8.45% -- 33.16%] | 41.44% [21.12% -- 61.76%] | 35.14% [25.36% -- 44.92%] | 50.04% [32.17% -- 67.92%] | 62.32% [53.67% -- 70.98%] |
|  | Any | One | Two | Three | Four or five |
| Scenario 1 | 2.21% [0.29% -- 4.13%] | -0.33% [3.31% -- -3.97%] | 4.79% [-2.33% -- 11.91%] | 9.17% [-5.55% -- 23.89%] | 4.93% [-23.75% -- 33.61%] |
| Scenario 2 | 9.81% [6.95% -- 12.67%] | 2.95% [-2.23% -- 8.13%] | 16.06% [6.64% -- 25.49%] | 26.36% [8.33% -- 44.39%] | 32.34% [-2.67% -- 67.35%] |
| Scenario 3 | 36.45% [31.82% -- 41.08%] | 19.54% [12.57% -- 26.51%] | 59.53% [48.43% -- 70.64%] | 57.18% [36.58% -- 77.77%] | 83.94% [47.17% -- 120.70%] |
| **Panel B Absolute reduction in adverse outcomes from three hypothetical levelling up scenarios** |  |  |  |  |  |
|  | Psychological distress | Poor health | Obese | Smoke | Poor academic achievement |
| Scenario 1 | 12 [-144 -- 169] | 65 [-75 -- 206] | 19 [-162 -- 200] | 113 [-63 -- 288] | 314 [106 -- 522] |
| Scenario 2 | 66 [-154 -- 285] | 208 [390 -- 27] | 254 [501 -- 6] | 357 [572 -- 141] | 1138 [1319 -- 957] |
| Scenario 3 | 465 [193 -- 737] | 483 [701 -- 265] | 1247 [1575 -- 920] | 793 [1045 -- 541] | 3513 [3880 -- 3145] |
|  | Any | One | Two | Three | Four or five |
| Scenario 1 | 200 [26 -- 373] | -17 [-211 -- 176] | 124 [-60 -- 307] | 81 [-49 -- 210] | 13 [-61 -- 86] |
| Scenario 2 | 886 [-155 -- 1926] | 157 [-118 -- 432] | 414 [171 -- 657] | 232 [73 -- 391] | 82 [-7 -- 172] |
| Scenario 3 | 3292 [2909 -- 3675] | 1039 [669 -- 1409] | 1535 [1249 -- 1821] | 504 [322 -- 685] | 214 [120 -- 308] |
| **Panel C Absolute reduction in adverse outcomes from three hypothetical levelling up scenarios (national level)** |  |  |  |  |  |
|  | Psychological distress | Poor health | Obese | Smoke | Poor academic achievement |
| Scenario 1 | 587 [-6848 -- 8022] | 3103 [-3573 -- 9778] | 913 [-7706 -- 9531] | 5370 [-2972 -- 13713] | 14941 [5057 -- 24825] |
| Scenario 2 | 3115 [-7307 -- 13537] | 9910 [18524 -- 1295] | 12055 [23805 -- 306] | 16961 [27200 -- 6722] | 54095 [62709 -- 45480] |
| Scenario 3 | 22121 [9189 -- 35054] | 22962 [33321 -- 12604] | 59304 [74876 -- 43733] | 37705 [49681 -- 25730] | 167010 [184493 -- 149527] |
|  | Any | One | Two | Three | Four or five |
| Scenario 1 | 9492 [1248 -- 17737] | -823 [-10025 -- 8379] | 5878 [-2850 -- 14606] | 3840 [-2327 -- 10007] | 597 [-2879 -- 4074] |
| Scenario 2 | 42122 [-7348 -- 91592] | 7466 [-5625 -- 20558] | 19693 [8138 -- 31248] | 11043 [3489 -- 18596] | 3920 [-323 -- 8164] |
| Scenario 3 | 156506 [138290 -- 174721] | 49398 [31786 -- 67010] | 72982 [59370 -- 86594] | 23951 [15325 -- 32578] | 10174 [5718 -- 14631] |
| Note: National level in Panel C is based on a UK population of 724,823 individuals aged 17 in 2018 when the fieldwork was carried out. |  |  |  |  |  |

**Table S10:**
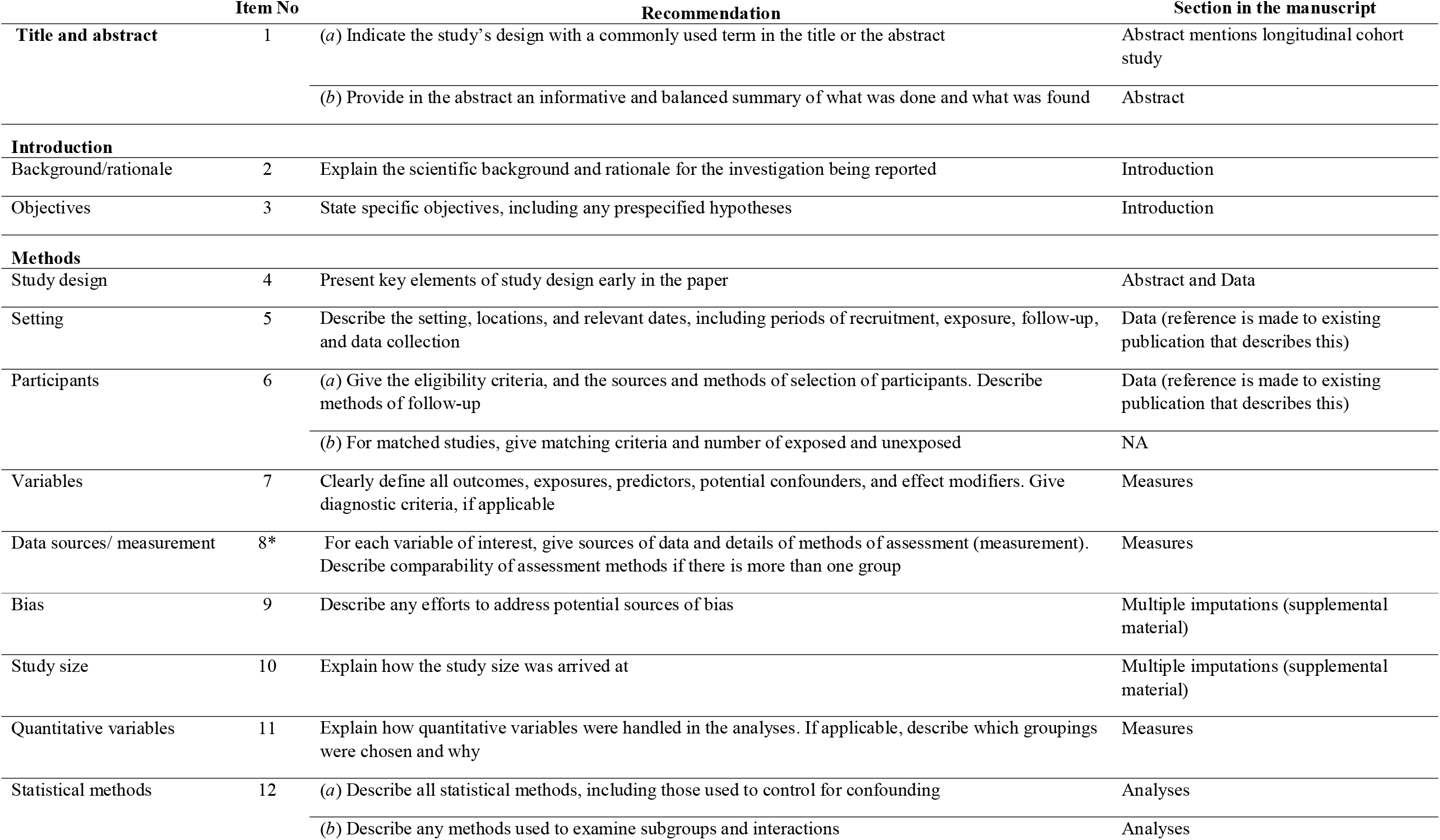
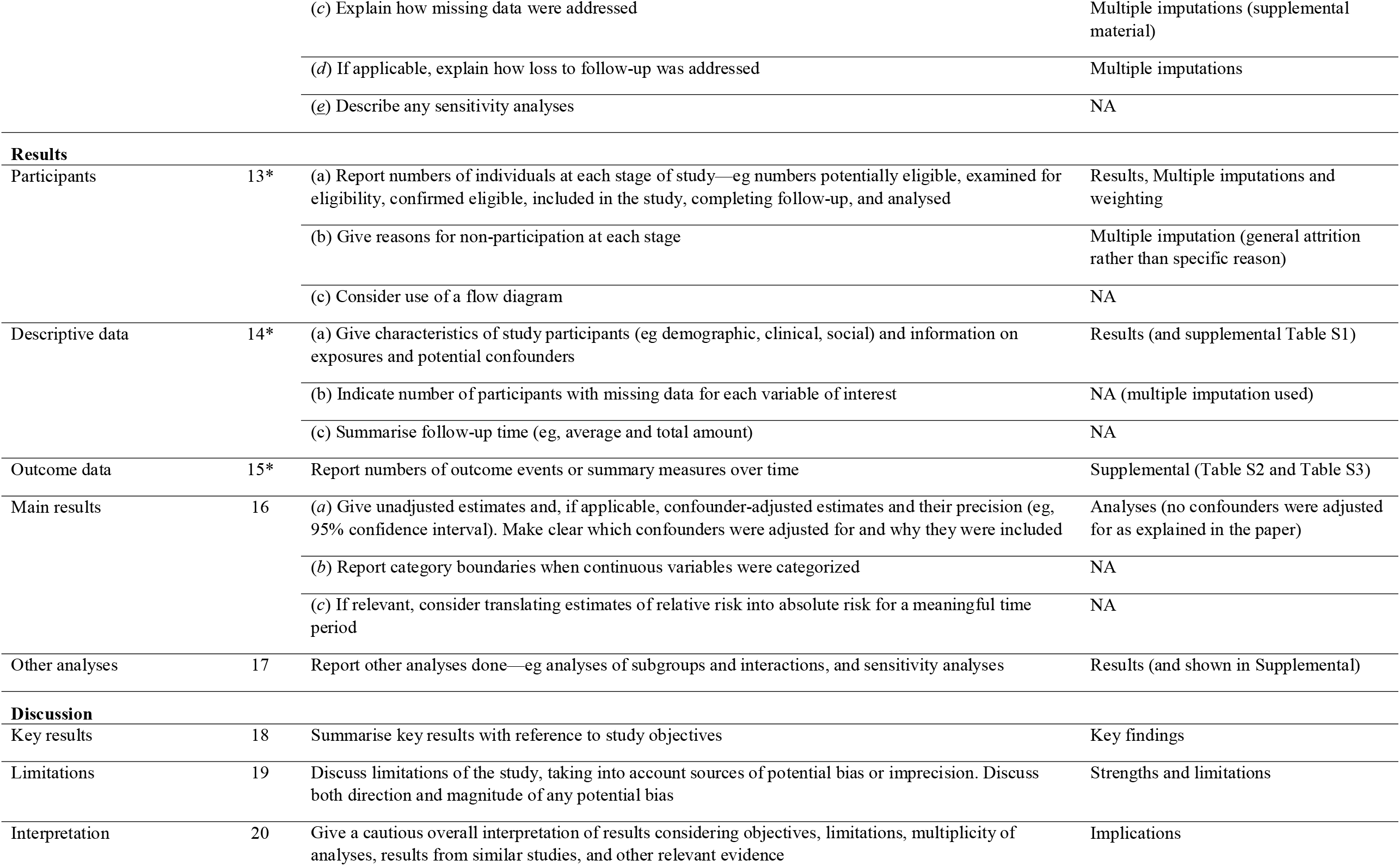
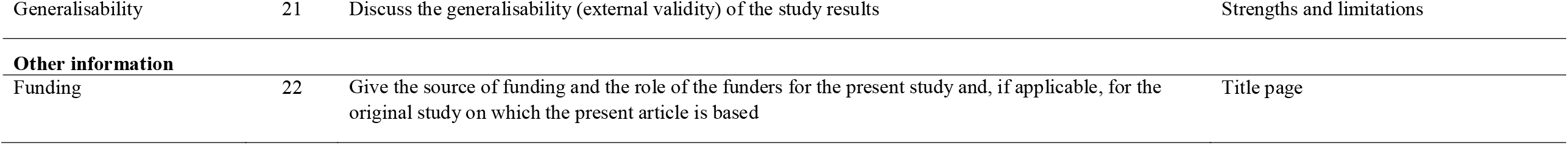
STROBE checklist (for cohort studies)

|  | Item No | Recommendation | Section in the manuscript |
| --- | --- | --- | --- |
| Title and abstract | 1 | (a) Indicate the study’s design with a commonly used term in the title or the abstract | Abstract mentions longitudinal cohort study |
|  |  | (b) Provide in the abstract an informative and balanced summary of what was done and what was found | Abstract |
| Introduction |  |  |  |
| Background/rationale | 2 | Explain the scientific background and rationale for the investigation being reported | Introduction |
| Objectives | 3 | State specific objectives, including any prespecified hypotheses | Introduction |
| Methods |  |  |  |
| Study design | 4 | Present key elements of study design early in the paper | Abstract and Data |
| Setting | 5 | Describe the setting, locations, and relevant dates, including periods of recruitment, exposure, follow-up, and data collection | Data (reference is made to existing publication that describes this) |
| Participants | 6 | (a) Give the eligibility criteria, and the sources and methods of selection of participants. Describe methods of follow-up | Data (reference is made to existing publication that describes this) |
|  |  | (b) For matched studies, give matching criteria and number of exposed and unexposed | NA |
| Variables | 7 | Clearly define all outcomes, exposures, predictors, potential confounders, and effect modifiers. Give diagnostic criteria, if applicable | Measures |
| Data sources/ measurement | 8* | For each variable of interest, give sources of data and details of methods of assessment (measurement). Describe comparability of assessment methods if there is more than one group | Measures |
| Bias | 9 | Describe any efforts to address potential sources of bias | Multiple imputations (supplemental material) |
| Study size | 10 | Explain how the study size was arrived at | Multiple imputations (supplemental material) |
| Quantitative variables | 11 | Explain how quantitative variables were handled in the analyses. If applicable, describe which groupings were chosen and why | Measures |
| Statistical methods | 12 | (a) Describe all statistical methods, including those used to control for confounding | Analyses |
|  |  | (b) Describe any methods used to examine subgroups and interactions | Analyses |
|  |  | (c) Explain how missing data were addressed | Multiple imputations (supplemental material) |
|  |  | (d) If applicable, explain how loss to follow-up was addressed | Multiple imputations |
|  |  | (e) Describe any sensitivity analyses | NA |
| **Results** |  |  |  |
| Participants | 13* | (a) Report numbers of individuals at each stage of study—eg numbers potentially eligible, examined for eligibility, confirmed eligible, included in the study, completing follow-up, and analysed | Results, Multiple imputations and weighting |
|  |  | (b) Give reasons for non-participation at each stage | Multiple imputation (general attrition rather than specific reason) |
|  |  | (c) Consider use of a flow diagram | NA |
| Descriptive data | 14* | (a) Give characteristics of study participants (eg demographic, clinical, social) and information on exposures and potential confounders | Results (and supplemental Table S1) |
|  |  | (b) Indicate number of participants with missing data for each variable of interest | NA (multiple imputation used) |
|  |  | (c) Summarise follow-up time (eg, average and total amount) | NA |
| Outcome data | 15* | Report numbers of outcome events or summary measures over time | Supplemental (Table S2 and Table S3) |
| Main results | 16 | (a) Give unadjusted estimates and, if applicable, confounder-adjusted estimates and their precision (eg, 95% confidence interval). Make clear which confounders were adjusted for and why they were included | Analyses (no confounders were adjusted for as explained in the paper) |
|  |  | (b) Report category boundaries when continuous variables were categorized | NA |
|  |  | (c) If relevant, consider translating estimates of relative risk into absolute risk for a meaningful time period | NA |
| Other analyses | 17 | Report other analyses done—eg analyses of subgroups and interactions, and sensitivity analyses | Results (and shown in Supplemental) |
| **Discussion** |  |  |  |
| Key results | 18 | Summarise key results with reference to study objectives | Key findings |
| Limitations | 19 | Discuss limitations of the study, taking into account sources of potential bias or imprecision. Discuss both direction and magnitude of any potential bias | Strengths and limitations |
| Interpretation | 20 | Give a cautious overall interpretation of results considering objectives, limitations, multiplicity of analyses, results from similar studies, and other relevant evidence | Implications |
| Generalisability | 21 | Discuss the generalisability (external validity) of the study results | Strengths and limitations |
| **Other information** |  |  |  |
| Funding | 22 | Give the source of funding and the role of the funders for the present study and, if applicable, for the original study on which the present article is based | Title page |

## Footnotes

a Mostafa T, Narayanan M, Pongiglione B, et al. Missing at random assumption made more plausible: evidence from the 1958 British birth cohort. Journal of Clinical Epidemiology 2021;136:44-54. doi: https://doi.org/10.1016/j.jclinepi.2021.02.019

b Mishra S, Khare D. On comparative performance of multiple imputation methods for moderate to large proportions of missing data in clinical trials: a simulation study. Med Stat Inform 2014;2(1):9. doi: doi: 10.7243/2053-7662-2-9

c Von Hippel P, Lynch J. Efficiency gains from using auxiliary variables in imputation. arXiv 2013;1311.5249 doi: https://doi.org/10.48550/arXiv.1311.5249

## Notes

### Competing Interest Statement

The authors have declared no competing interest.

### Funding Statement

This study was funded by UK Prevention Research Partnership (MR/S037527/1)

### Author Declarations

The data used are openly available data secondary data available from the UK Data Service. Series: beta.ukdataservice.ac.uk/datacatalogue/series/series?id=2000031

